# Intersex Exceptions in U.S. Laws Restricting Medical Care for Transgender Minors

**DOI:** 10.1101/2025.04.22.25326142

**Authors:** Sara A. Mar, Gnendy Indig, Meredithe McNamara, Elizabeth R. Boskey, Katharine B. Dalke, Elizabeth Dietz, Morgan Carpenter, Cynthia Kraus, Erika Lorshbough, Quinnehtukqut McLamore, Kimberly Zieselman, Brian D. Earp, Hannah C. Wenger

## Abstract

Contemporary U.S. legislation restricts hormonal and surgical interventions for transgender and gender-diverse (TGD) persons, primarily legal minors, who may need them and who voluntarily request them. We conducted a comprehensive review of all U.S. legislation from 2021 to 2024 that restricts voluntarily sought TGD-related healthcare and found explicit allowances for physically comparable, but non-voluntary interventions to “normalize” the benign bodily attributes of children with intersex conditions, or congenital variations in sex characteristics. Our findings could suggest a lack of equal regard for all individuals’ moral and legal rights to self-determination and bodily autonomy.

## INTRODUCTION

Since 2021, 27 U.S. states have enacted legal restrictions, or bans, on voluntary medical interventions (commonly termed gender-affirming medical care or GAMC) for transgender and gender diverse (TGD) individuals, primarily legal minors (Appendix). Supporters of these bans have argued that the restrictions are necessary to protect TGD minors from potentially risky medical interventions that alter benign sexual anatomy – that is, non-diseased anatomy that does not, in and of itself, threaten the health of the individual – and allege that the minors are too young to understand or consent to such interventions, notwithstanding parental permission.

Seldom acknowledged in the sociopolitical discourse about these bans are their statutory exceptions, or carve-outs, that specifically exempt – and implicitly endorse – non-voluntary, appearance-“normalizing” interventions on the benign bodily attributes of presumptively non-TGD individuals, primarily those with variations of sex characteristics (VSCs) or intersex traits (discussed further below). These interventions affect similar anatomy to GAMC, carry analogous physical risks, and are of unknown benefit. Yet, unlike GAMC for TGD minors, they are regularly performed before the child is able to participate meaningfully in the decision-making process regarding their own body.

To elucidate this tension, we conducted a comprehensive review and analysis of all laws between 2021 and 2024 that limit GAMC for TGD populations in the United States, paying special attention to how sex and gender are defined in the laws and to the justifications, if any, provided for intersex-related or other exceptions to the prohibitions on GAMC. Our methodology and findings are highlighted below with full details and documentation including search strategy located in the Appendix.

### Background

To frame our study, we begin with a brief overview of intersex and TGD populations and how they differ in certain ways, while also having some overlapping medical needs. Sex and gender are often oversimplified and inaccurately regarded as being synonymous, sometimes leading to a conflation of intersex and TGD concepts (see Appendix Table 1 for definitions). Despite considerable political contestation over the nature and implications of these categories within social and legal spheres, the complexities of sex and gender – and the experiences of persons living outside of traditional sex or gender classifications – are increasingly salient within medicine and the biological sciences.(1–5)

At birth, sex is usually recorded as female or male following visual inspection of an infant’s genitalia. For infants born with what are variously called differences of sex development, intersex traits, or congenital variations in sex characteristics (VSCs),(6,7) their anatomical features, hormone profiles, or karyotype are judged to differ from normative biomedical criteria for the female or male sex. Though recent and reliable prevalence data are unavailable for the intersex population, it is estimated that a fraction of 1% to under 2% of babies are born with VSCs.(8–13) In some such cases, additional medical tests may be performed before a decision is made about sex classification. Alongside biological information, this decision may be informed by social or cultural beliefs, such as perceived ideals regarding femininity or masculinity,(14–17) which may or may not align with the individual’s future identity or values.

Among more than 30 VSCs and at least 100 genes linked to development of diverse sex characteristics,(12) some conditions may be associated with serious physical health risks that necessitate urgent medical intervention (see endnote 18). However, most VSCs do not portend significant physical impairment or risk of death, and any associated diverse-appearing genitalia are not inherently pathological. Still, hormonal and surgical interventions are often performed on infants and children born with benign or low-risk VSCs in an effort to conform their bodies to what is considered typical for females or males. Mostly, this is done on a non-voluntary basis before the child can consent or assent. This preempts opportunities for individuals born with VSCs, some of whom may later identify as intersex, to freely develop and express their own values and preferences about their bodies; to pursue the option of leaving their sexual or reproductive anatomy unmodified, or differently modified; and, if modification is judged desirable, to be personally informed of the risks of such “normalizing” procedures so as to weigh these against any potential benefits.(19–21)

Most infants are reared as if their birth-recorded sex accurately describes both their internal and external sex characteristics and their self-conception or experienced identity in relation to sex or gender, sometimes termed “gender identity.”(22) Most people living in the U.S., whether intersex or non-intersex (i.e., “endosex”)(23), have a gender identity that aligns with what is culturally prescribed for individuals with their birth-recorded sex. Such individuals are often termed cisgender or non-transgender (non-TGD). Approximately 1% experience a gender identity that differs from what is normatively associated with their birth-recorded sex.(24,25) These individuals may be described, or may self-identify, as transgender or gender-diverse (TGD).

Gender-affirming medical care (GAMC) refers to interventions that are intended to resolve felt discrepancies between physical embodiment and gender identity. These discrepancies can be highly distressing and can have a significantly negative impact on well-being if left unaddressed. For TGD individuals, GAMC may consist of hormonal and non-hormonal medications, vocal training, mental health support, fertility treatments, and/or surgeries, among other modalities. Importantly, GAMC is a common healthcare need for both TGD and non-TGD individuals (e.g., interventions to address gynecomastia in non-TGD males; see endnote 26).(27–29)

Irrespective of whether a person has been medically classified as female or male (with or without the presence of VSCs), or identifies as intersex, TGD, or otherwise, individuals of all body types and identities ought to have the equal opportunity to pursue, or to forgo, GAMC or other similar interventions on a voluntary basis, in accordance with the fundamental human rights to non-discrimination, self-determination, and bodily autonomy.(19,20) However, legal and political processes may affect this equality of opportunity, as shown herein.(30–33) We searched and analyzed U.S. legislation from 2021 to 2024 that prohibits voluntarily sought TGD-related healthcare and found explicit allowances for physically comparable, but non-voluntary interventions to “normalize” the benign bodily attributes of children with intersex conditions, or VSCs. As a group with practical and scholarly expertise in the fields of medicine, law, and ethics, we considered legal and ethical aspects of how U.S. jurisdictions statutorily define sex and gender and treat healthcare services that alter sexual anatomy, with a focus on categorical restrictions and exceptions.

## METHODOLOGY

We searched for statutes across four full legislative sessions (January 1, 2021, to December 31, 2024) that restrict healthcare services for TGD individuals in all 50 U.S. states, the District of Columbia, U.S. territories, and tribal and federal jurisdictions (see Appendix Box 1 and Figure 1 for detailed search methodology).(36) Statutes were assessed through sequential, independent review by two authors (SAM, HCW). For each statute, the authors identified descriptions of sex, gender, and GAMC; restricted healthcare services; stated purpose(s) for which services are restricted; exceptions to restrictions; and penalties for statutory violations. All quoted text denotes statutory language.

**Figure 1.**
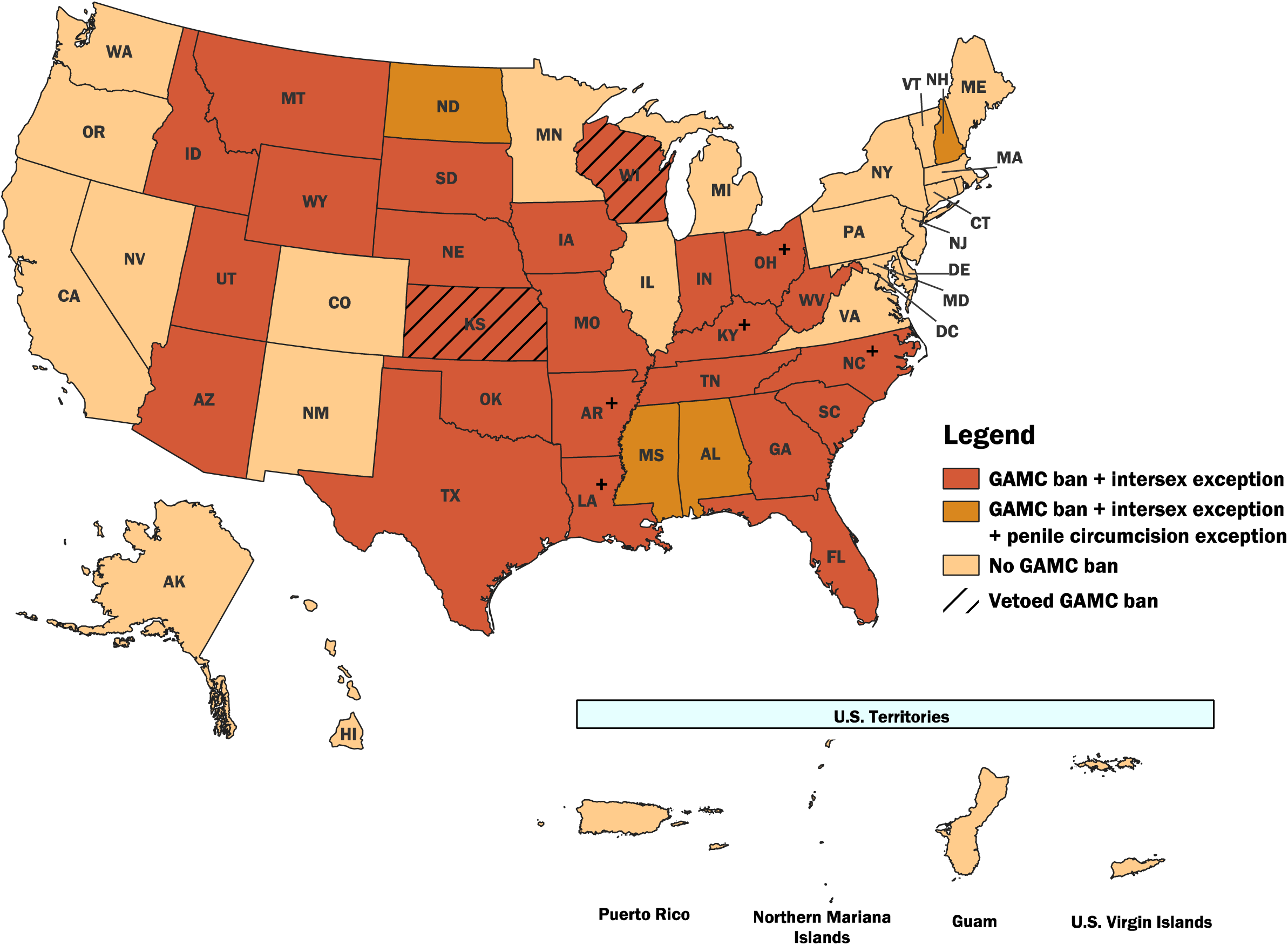
Map of intersex and penile circumcision exceptions in U.S. laws* restricting medical care for transgender minors, 2021-2024. GAMC = Gender-Affirming Medical Care. No GAMC bans were identified in the District of Columbia, U.S. territories, or tribal and federal jurisdictions during the study period. (+) denotes a state in which a GAMC ban was enacted after a veto override.

## RESULTS

We identified 28 U.S. states with a combined total of 30 passed or enacted statutes (i.e., bans) that restrict healthcare services for TGD minors (Appendix Table 2), including 2 states (KS, TN) that passed a ban in 2 separate legislative sessions.(37) Three of the 30 bans were passed by the legislature but permanently vetoed by the governor (KS, WI), leaving 26 states that enacted a ban in the study period.(38) No bans were identified in the District of Columbia, U.S. territories, or tribal and federal jurisdictions. Twenty-eight of the identified 30 bans prohibit, for the purposes of “gender transition” (or a similar term), the use of medications that suspend pubertal development and/or the use of hormone therapies. Twenty-nine bans prohibit genital and/or non-genital surgeries. Fourteen bans also mention that medical and surgical care is disallowed if it affects or removes a minor’s “healthy,” “functional,” or “nondiseased” body parts, including characteristics “typical” for their biological sex. None specifies the party responsible for making such determinations.

All 30 bans denote statutory exceptions, or instances in which restricted care is permitted (Figure 1 below; Appendix Tables 2-3). Twenty-nine of the 30 bans contain an exception for interventions on individuals with VSCs; Tennessee’s 2021 ban does not include such an exception, but this statute was superseded by a 2023 ban that does.(39,40) Some otherwise restricted care may be allowed under 20 bans for undefined “infection, injury, disease, or disorder…caused by or exacerbated by [GAMC],” and under 16 bans for unspecified physical conditions that place an individual at risk for “death or impairment of major bodily function.” West Virginia’s 2023 ban permits GAMC for so-called “severe” gender dysphoria, situating it as the only state that does not have an absolute TGD-specific prohibition. Four bans include an exception for penile circumcision. Additional exceptions are cataloged in Appendix Table 3.

Twenty-four of the 30 bans define sex as a binary biological condition of being female or male, as determined by chromosomes, endogenous steroid hormones, reproductive potential, internal reproductive organs, and/or external genitalia present at birth (Appendix Table 2). Sixteen bans invoke the concept of “nonambiguous” genitalia as a criterion for their definition. Twelve bans define gender as “the psychological, behavioral, social and cultural aspects of being male or female,” without mention of TGD identities. Eleven bans describe GAMC as a form of “gender transition” or a similar term. Referring a TGD minor for – or otherwise “aid[ing] and abet[ting]” – restricted GAMC is prohibited in seven bans. Punitive actions vary across bans and may involve loss of licensure, financial penalties, and criminal and civil charges for healthcare professionals (HCPs).

## DISCUSSION

GAMC bans prohibit hormonal or surgical alterations to the benign sexual anatomy of TGD minors. This applies even to adolescents who may both need and want this care within their states of residence, and who are supported in accessing it on a voluntary basis by their parents and HCPs. However, statutory carve-outs for intersex “normalization” procedures explicitly permit such interventions even when non-voluntary, raising questions about potential double standards.

Interventions on intersex individuals are not the only excepted circumstances involving benign sexual anatomy within the bans. In the United States, uniquely among high-income countries, non-therapeutic, non-religious (so-called “routine”) infant penile circumcision remains a majority birth custom. Like intersex “normalization” procedures, such circumcision is performed in healthcare settings without the consent or assent of the individual and in the absence of a physical health condition that requires urgent medical intervention.(41) Several bans explicitly legitimize such circumcisions on healthy endosex male infants, such as New Hampshire’s statute permitting HCPs to remove benign penile foreskin for “religious, cultural or health reasons.”(42) Beyond circumcision, labiaplasty(43) and other cosmetic surgeries that alter or remove non-diseased sexual or genital tissues are increasingly performed on non-TGD minors(44–47) and seem set to continue in states with GAMC bans, although they are neither explicitly restricted nor excepted.

### Carve-Outs Undermine Arguments for GAMC Restrictions

Proponents of the GAMC bans who also support the carve-outs for intersex interventions might argue that, unlike children without VSCs (inclusive of TGD minors), children born with VSCs have a recognized physical health condition and are therefore appropriate subjects of surgical or hormonal medical treatment, even without their personal consent or assent. However, as noted, most VSCs do not pose a significant or time-sensitive threat to physical health, which undermines the force of this position. Such a justification also fails to explain the carve-outs for non-therapeutic infant penile circumcision.(49) Like intersex “normalization” procedures, “routine” circumcision in the U.S. serves to conform a child’s body to what is considered typical – or culturally acceptable – for members of one’s designated sex class and presumed gender identity, irrespective of consent or medical necessity.(50) This suggests that the carve-outs, taken together, may have more to do with upholding traditional gender norms about bodily appearance than with addressing a relevant physical health concern.(50)

Proponents of GAMC bans argue that there is scientific uncertainty about the effectiveness of GAMC interventions in bringing about positive outcomes. The most oft-cited review in this area(51) characterizes the existing evidence base as being generally of a low quality. This review has itself also been subject to critique.(52–54) However, such discourse by ban proponents – about the ostensibly low quality of research for GAMC – ignores the fact that the evidence base regarding long-term causal benefits of non-voluntary intersex procedures (compared to no, or voluntary procedures) is of no greater, or arguably even lower, quality.(19)

Of course, better-quality research is desirable in all areas of healthcare, particularly when dealing with vulnerable populations. The point here is simply one of consistency: if the quality of evidence supporting one set of interventions is deemed, by some, to be too low to permit even case-by-case decision-making by HCPs, parents, and older minors who may seek such interventions, then similar or even lower-quality evidence supporting analogous, but non-voluntary interventions in younger minors should inspire, if anything, even greater concern. And yet, the opposite pattern of concern is what is suggested by the bans, in terms of what is permitted versus restricted.

Intersex procedures are typically performed at an early age, when the child’s future (i.e., adolescent or adult) gender identity – and any associated bodily preferences or health needs they may have at that point – are a matter of speculation. By contrast, for TGD individuals who actively request GAMC in adolescence or adulthood, such information is already available and can be factored into a shared decision-making process around potential medical interventions, with the meaningful participation of the individual concerned. This allows for judgments to be made on a case-by-case basis with due consideration for the young person’s known (rather than merely anticipated, future) health or identity-related needs and circumstances.(55,56) Thus, the ubiquitous intersex carve-out within the GAMC bans may signal a desire to reinforce traditional standards of what is considered “normal” in relation to sex or gender, irrespective of voluntariness, rather than to make a principled distinction based on relative risks and benefits or a minor’s capacity to consent.

### Narrow Sex and Gender Definitions

In the GAMC bans, both intersex people and TGD people are excluded from the narrow definitions of sex and gender that are employed, respectively. The bans criminalize HCPs who provide or refer for voluntary GAMC for TGD minors, while codifying practices that allow similar, but non-voluntary treatments on the sexual anatomy of intersex minors. Infertility, sexual dysfunction, and other physical health risks inherent to medical interventions on benign sexual anatomy are, paradoxically, proffered to justify restrictions on care for TGD minors but disregarded within statutory carve-outs for similar procedures on non-TGD minors.

At stake is the health, well-being, and equal rights not only of TGD individuals, but also of non-TGD individuals, whether or not they have intersex traits. The right of all individuals to the highest standard of healthcare, which includes respect for self-determination and bodily autonomy, is threatened by contradictory laws that defer to the judgment of parents and HCPs in some instances (i.e., for non-TGD minors) while superseding them in others (i.e., for TGD minors). Such legislation is antithetical to increasing global calls to develop consistent legal and ethical protections that center the preferences and values of young people in decision-making about medical interventions that affect their own genital and sexual anatomy.(19,20,57,58)

Whether TGD or non-TGD, intersex or endosex, all individuals have equal right to make informed decisions about their bodies.(19,59) Ideally, any treatments for children with intersex traits should be offered within a multidisciplinary care model that prioritizes informed personal consent as an intrinsic part of so-called “patient centered” care. This approach makes psychosocial, psychotherapeutic, and peer supports accessible to the individual and their family, and ensures respect for the individual’s self-determination and bodily autonomy by obtaining age-appropriate consent or assent for any surgery or hormone therapies that may ultimately be pursued.(60–63) The same standard should apply to TGD and non-TGD minors.

### Relevance to the Politico-Legal Landscape in the United States and Beyond

Our findings elucidate the differential statutory rules applied to TGD and presumptively non-TGD minors, rules shaped by definitions of sex and gender that conflict with more nuanced scientific considerations of these concepts. Such legislation discriminately limits healthcare access and delivery for some, while endorsing medically unnecessary, non-voluntary care for others. Similarities across statutes (e.g., preambles, definitions, carve-outs) may reflect the influence of model legislation developed by medico-legal groups with particular views on sex, gender, and GAMC.(64,65)

Notably, state governors exercised veto powers for seven bans (though five vetoes were overridden), criticizing governmental overreach and highlighting potential unconstitutionality.(66–72) The U.S. Supreme Court is considering whether Tennessee’s ban on GAMC constitutes sex-based discrimination,(73) which is prohibited under the Equal Protection Clause of the U.S. Constitution’s 14th Amendment. Federal actions under the newly inaugurated Trump-Vance administration appear poised to perpetuate a comparable contradiction to that found in our analysis.

Importantly, while enacted GAMC bans in the U.S. may seem to permit appearance-“normalizing” alterations on infants and children born with VSCs, these interventions have neither been declared clearly legal or illegal under U.S. law. In contrast, numerous other countries have explicitly prohibited such practices.(74) The United Nations, World Health Organization, and U.S. Department of Health and Human Services under the Biden-Harris administration have also expressly endorsed the fundamental rights of people with intersex traits – like people of all sexes and genders – to bodily integrity and bodily autonomy.(7,75–77)

The Trump-Vance administration, however, is charting a federal policy agenda that, like the state legislation identified in our analysis, focuses on sex, gender, and GAMC. During the first two weeks of the current administration, an executive order (EO) established, for all federal policy and administrative purposes, a binary definition of sex based on the capacity to produce gametes at conception.(78) This theoretical definition is inconsistent with real-world methods of human sex classification, especially in the case of persons born with VSCs.(79) Gamete production is not assessed at conception or birth, and decisions made on the basis of actual or hypothetical gamete-making capacity may conflict with more comprehensive methods of classifying an individual’s sex based on a wider set of relevant features.(80,81)

Another EO defined GAMC as “chemical and surgical mutilation” of TGD individuals under the age of 19 years and called to prohibit federal activities that “fund, sponsor, promote, assist or support the so-called ‘transition’ of a child from one sex to another.”(82) Poised to variably impact GAMC for not only TGD minors, but also some TGD legal adults within healthcare systems across the United States, this executive communication has spawned litigation attempting to block its implementation.(83–87)

The EO also called for enforcement of the U.S. federal Female Genital Mutilation/Cutting (FGM/C) statute,(88) conflating GAMC with FGM/C despite ethically relevant differences. Notably, the U.S. federal prohibition on FGM/C forbids all medically unnecessary alterations to the external female genitalia of persons under the age of 18 years,(89) but has not been interpreted as protecting individuals born with VSCs from such alterations, even if they were classified as female at birth and if similar anatomical features (e.g., clitoral tissues) would be affected.(90–92) International FGM/C legislation also contains explicit or de facto intersex exceptions.(93,94)

While the enforceability and implementation of the EOs are uncertain, these federal moves implicate both TGD and intersex persons. Alongside state and federally(95,96) legislated restrictions on GAMC, the Trump-Vance administration has also orchestrated removal of peer-reviewed and other scientific information on TGD and intersex populations.(97)

### Limitations

This analysis excludes statutes passed or enacted during the 2025 legislative session, which is still in process as of publication. In February 2025, Kansas’ legislature overrode a gubernatorial veto to become the 27^th^ U.S. state to enact a GAMC ban.(98,99) In April 2025, the West Virginia legislature amended its 2023 ban,(100) notable for allowing GAMC for so-called “severe” gender dysphoria, to completely remove this exception (the revised law is awaiting the governor’s signature at the time of writing); in any case, this carveout is likely inoperable because gender dysphoria is not graded by severity in clinical practice (48). These 2025 bans contain equivalent definitions of sex and gender to those documented here (Appendix Table 2), alongside similar restrictions on hormonal and surgical interventions for TGD minors; exceptions for interventions on individuals with VSCs; and penalties for statutory violations.

## CONCLUSION

Legislative bans on GAMC across U.S. states restrict medical services that affect benign sexual anatomy, and do so in a discriminatory manner. Access to and delivery of healthcare services are limited for TGD individuals who desire them, while implicit permissions are codified for medically unnecessary, appearance-”normalizing” interventions on infants and children with VSCs, thus preempting their ability to personally consider, much less assent or consent to, such procedures. We conclude that neither TGD restrictions nor intersex exceptions within these bans protect minors’ right to self-determination and their growing capacities for bodily autonomy. They instead reinforce the pathologization of gender and anatomical diversity in the name of social norms and conformity. Sound health policy uses science and ethics to afford equal access to healthcare services without discrimination based on sex, gender, or other protected characteristics. Lawmakers and politicians across all jurisdictions in the United States must protect the moral and legal right of all individuals to seek or forgo care that modifies their sexual anatomy in a way that acknowledges and respects their bodily sovereignty.

## Data Availability

All data are available in the included appendix.

## APPENDIX

**Appendix Table 1.** Definitions for terminology used in this paper, conforming to broadly accepted standards within scientific and medical communities*.

| Term | Definition |
| --- | --- |
| Sex | In the biological sciences, sex typically refers to a classification of members of sexually reproducing species based on prototypical body plans and developmental trajectories associated with production of large or small gametes, respectively (i.e., eggs, sperm). In humans, formal sex categorization of individuals is typically carried out at birth, most routinely based on visible genital anatomy or other observed or inferred biological factors. In Western societies with modern healthcare systems, almost all individuals with congenital variations in sex characteristics (VSCs), sometimes called intersex traits, are designated by healthcare professionals as either female or male based on a biomedical-and-social interpretation of their internal and external sexual and reproductive features, taken together. However, in some cases, such a “binary” designation may not be clearly fitting or may otherwise be judged challenging to make. Moreover, such designations may or may not align with the individual’s own understanding or experience of their sex or gender later in life. |
| Endosex | Innate physical sex characteristics judged to fall within the broad range of what is considered typical or normal for binary female or male bodies according to standard biomedical criteria; or persons with such characteristics. |
| Intersex/Variations in Sex Characteristics (VSCs) | Sex traits or reproductive features that differ from normative biomedical criteria for typical female or male anatomy, characteristics that may be visible at the time of birth or present later in life. Persons with intersex traits or variations in sex characteristics (VSCs) are sometimes referred to, or may self-identify as, intersex, with or without associated binary sex terms (e.g., intersex person versus intersex man, intersex woman). However, many individuals with such traits regard themselves as either female or male, albeit with one or more disorders/differences in sex development (DSD/dsd). Common intersex/VSC conditions include gonadal dysgenesis, congenital adrenal hyperplasia, androgen insensitivity syndrome, and Turner syndrome. Penile hypospadias is sometimes included under the umbrella of intersex/VSCs. |
| Gender Identity | An individual’s self-conception or personal identity in relation to sex or gender, or their deeply ingrained sense of being a girl/woman, boy/man, or potentially another gender. May include physical, psychological, social, and cultural aspects. |
| Transgender and Gender-Diverse (TGD) | An individual whose gender identity is markedly different from what is culturally expected in their society based on their sex as designated at birth. |
| Non-Transgender (Cisgender) | An individual whose gender identity is not markedly different from what is culturally expected in their society based on their sex as designated at birth. |
| Gender-Affirming Medical Care (GAMC) | Health care that seeks to bring a person’s physical embodiment and behavior into felt alignment with their gender identity, or to resolve felt discrepancies (which may be highly distressing) between the same. Most commonly describes hormonal and surgical treatments for TGD individuals, but also applicable to non-TGD individuals. |

Sources: Carpenter M, Dalke KB, Earp BD. Endosex. J Med Ethics. 2023 Mar;49(3):225–6; Goymann W, Brumm H, Kappeler, PM. Biological sex is binary, even though there is a rainbow of sex roles. BioEssays. 2023; 45(2):e2200173; however, see Karkaziz, K. The misuses of “biological sex.” Lancet. 2019;394(10212):1898-1899. Notes: \*May differ from statutory definitions within this paper

**Appendix Table 2.**
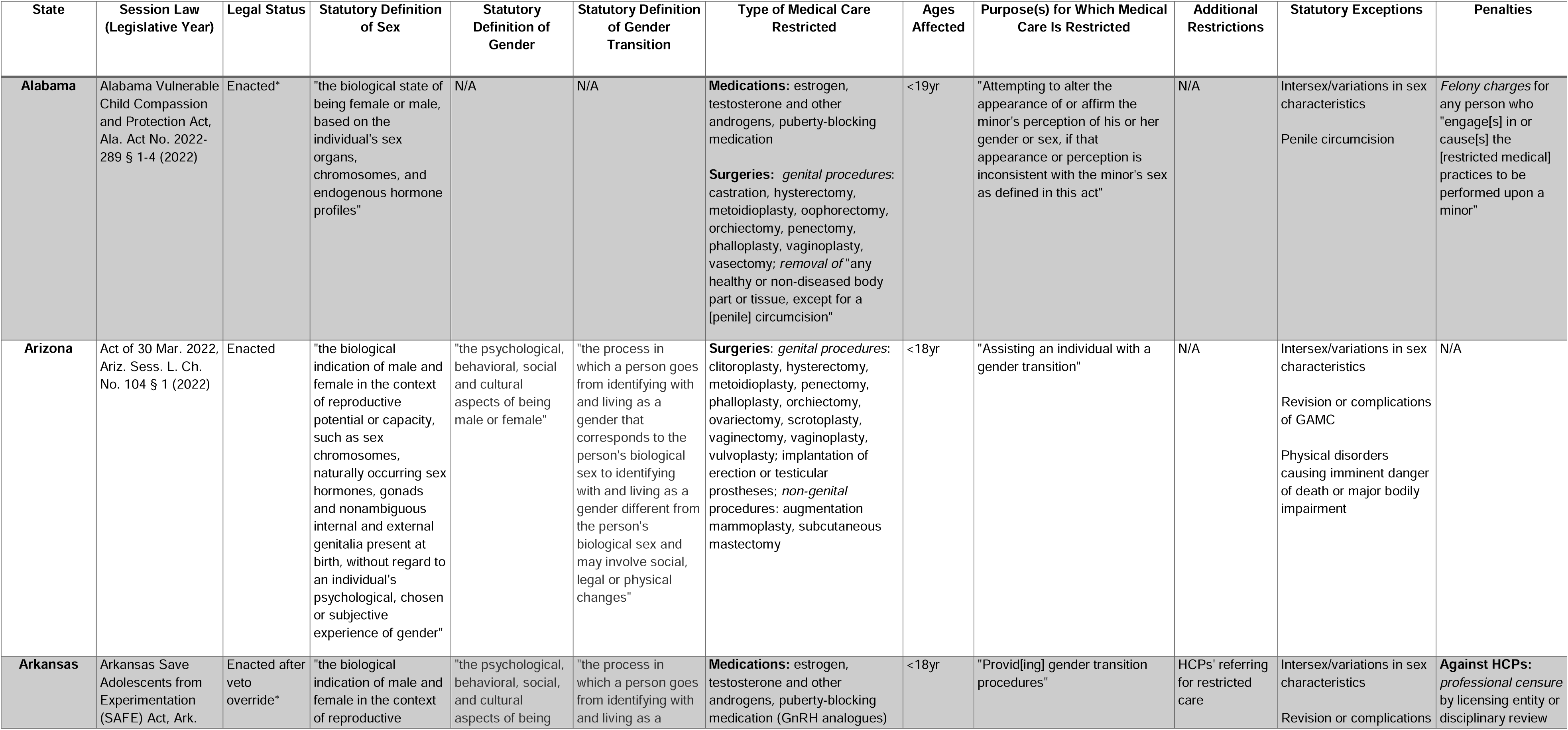
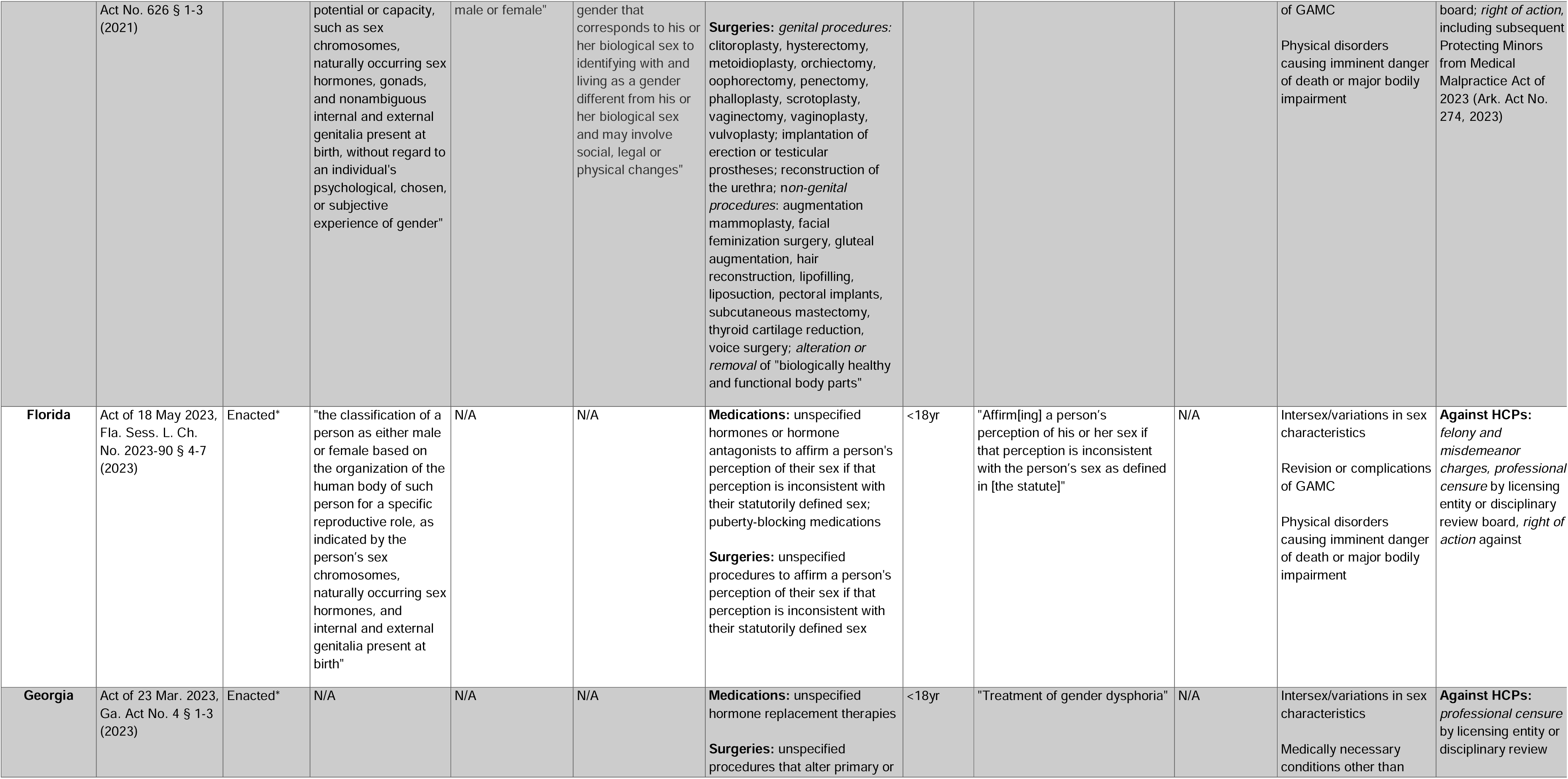
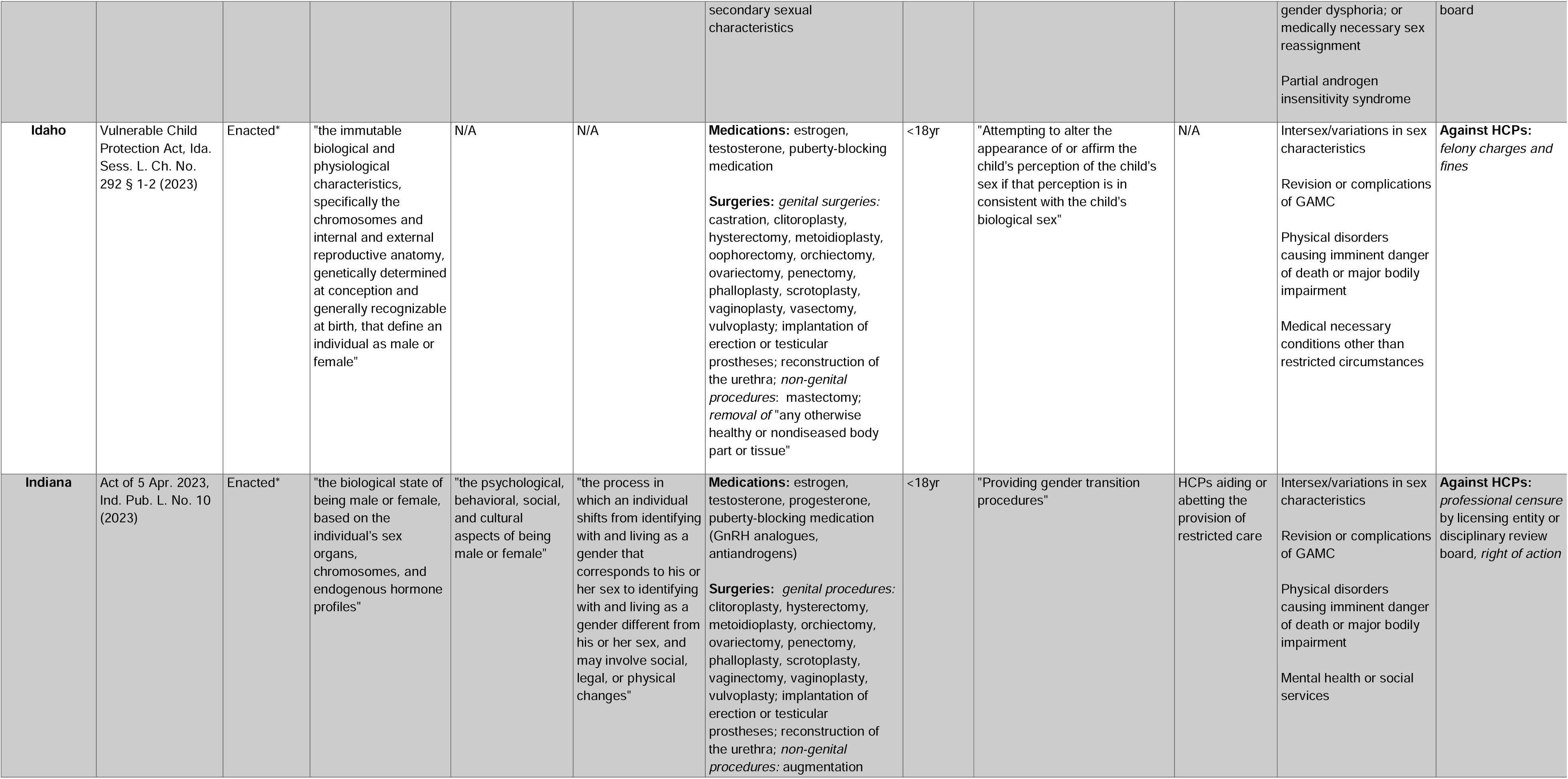
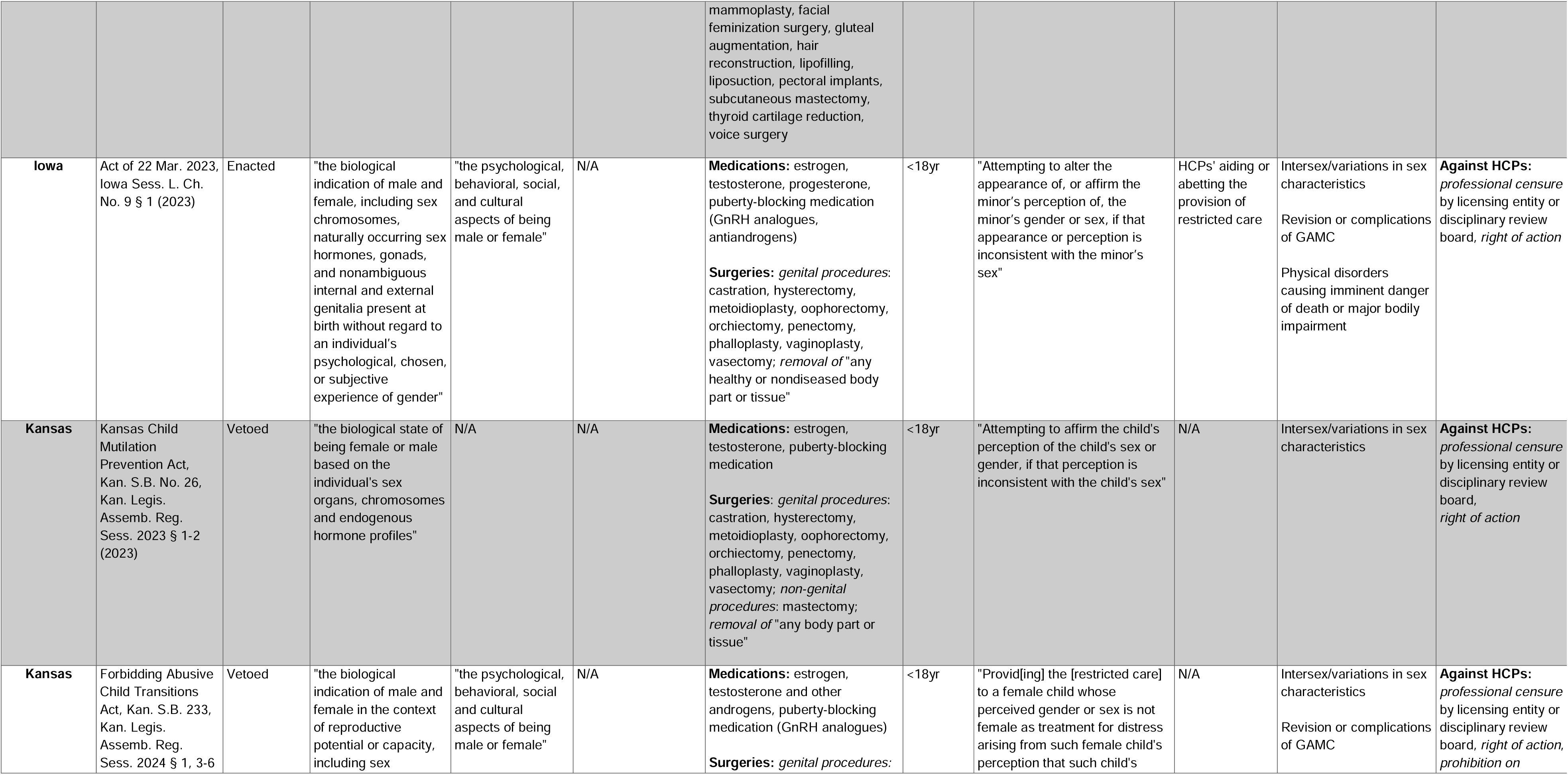
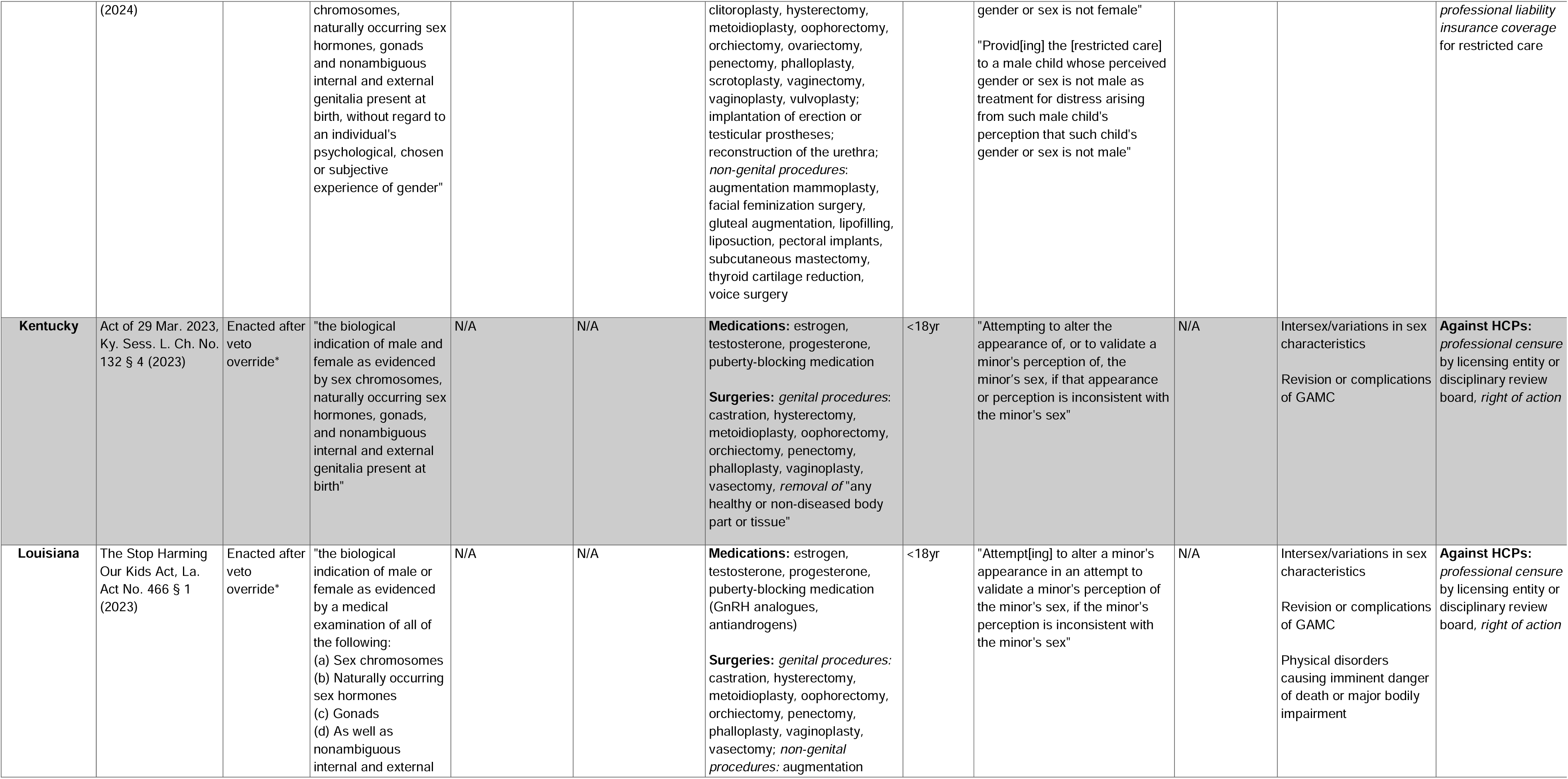
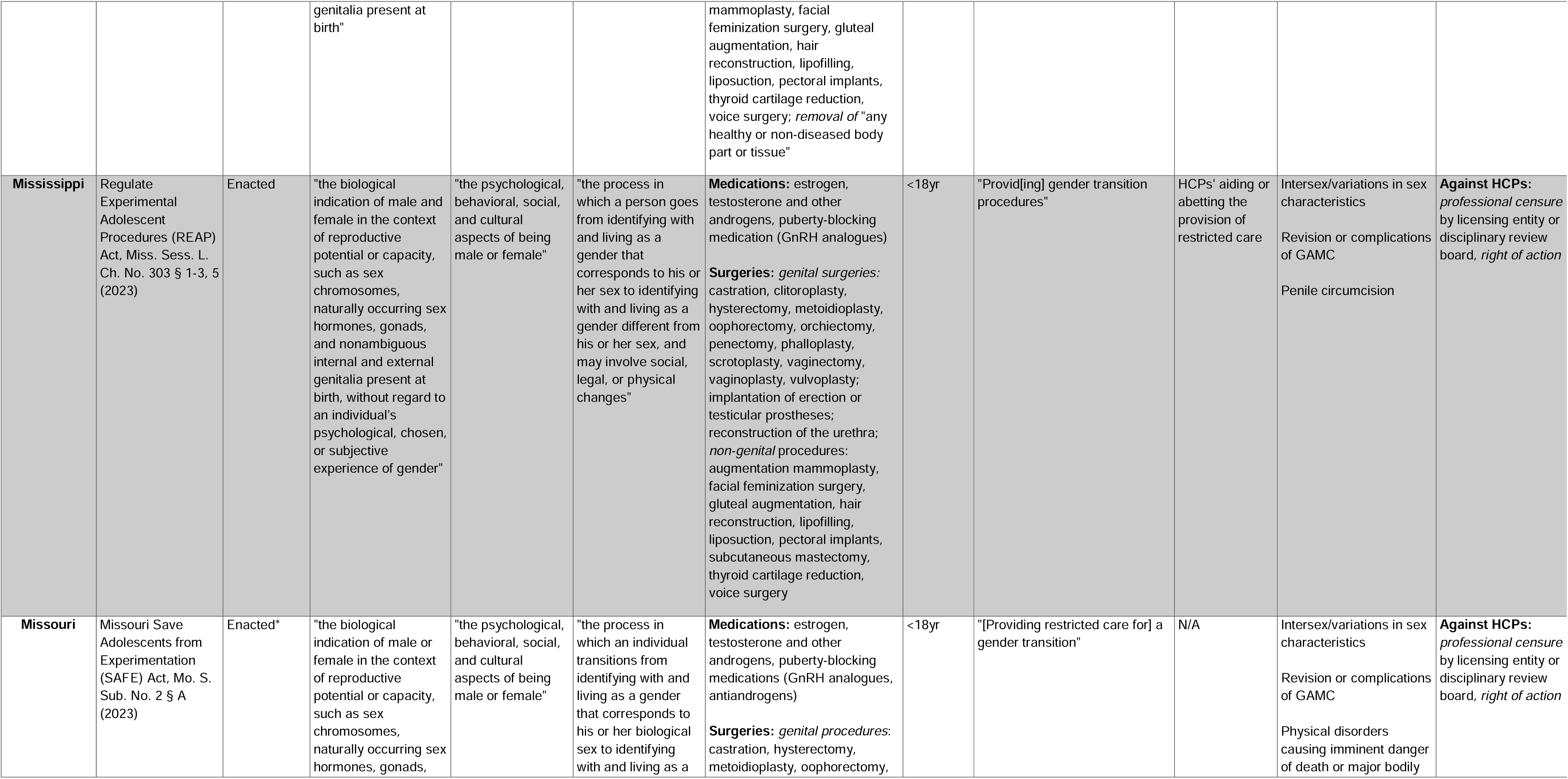
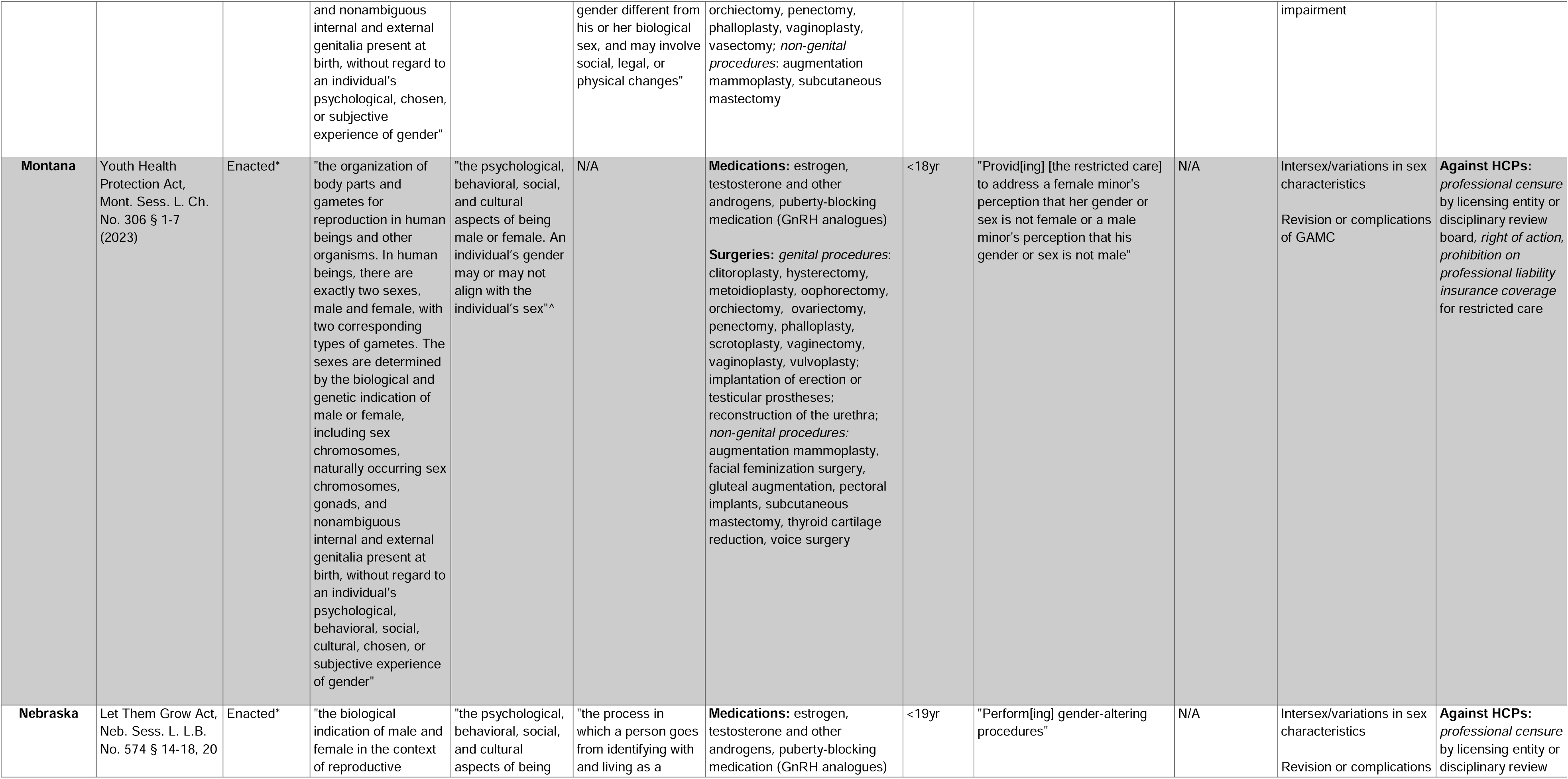
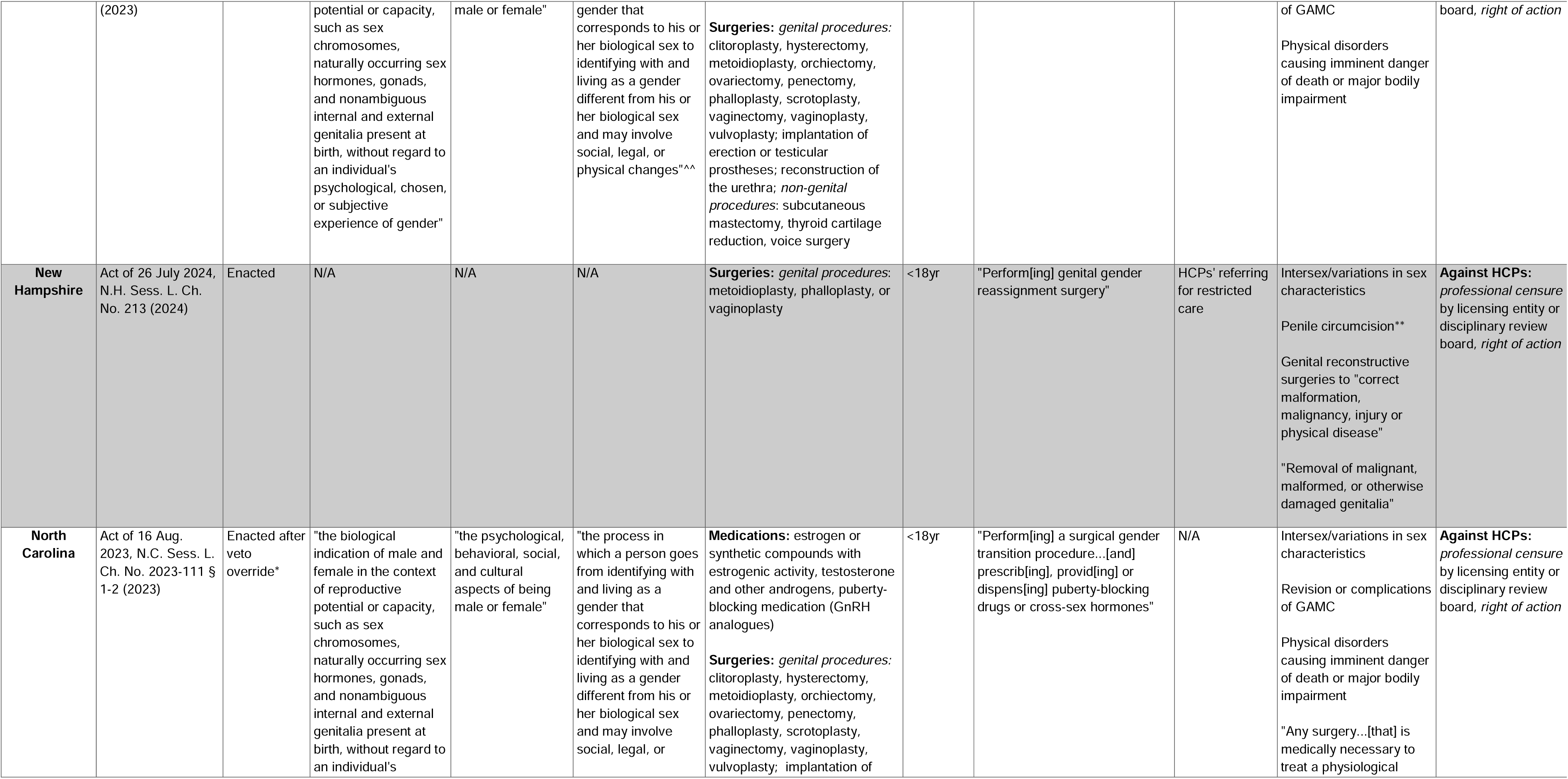
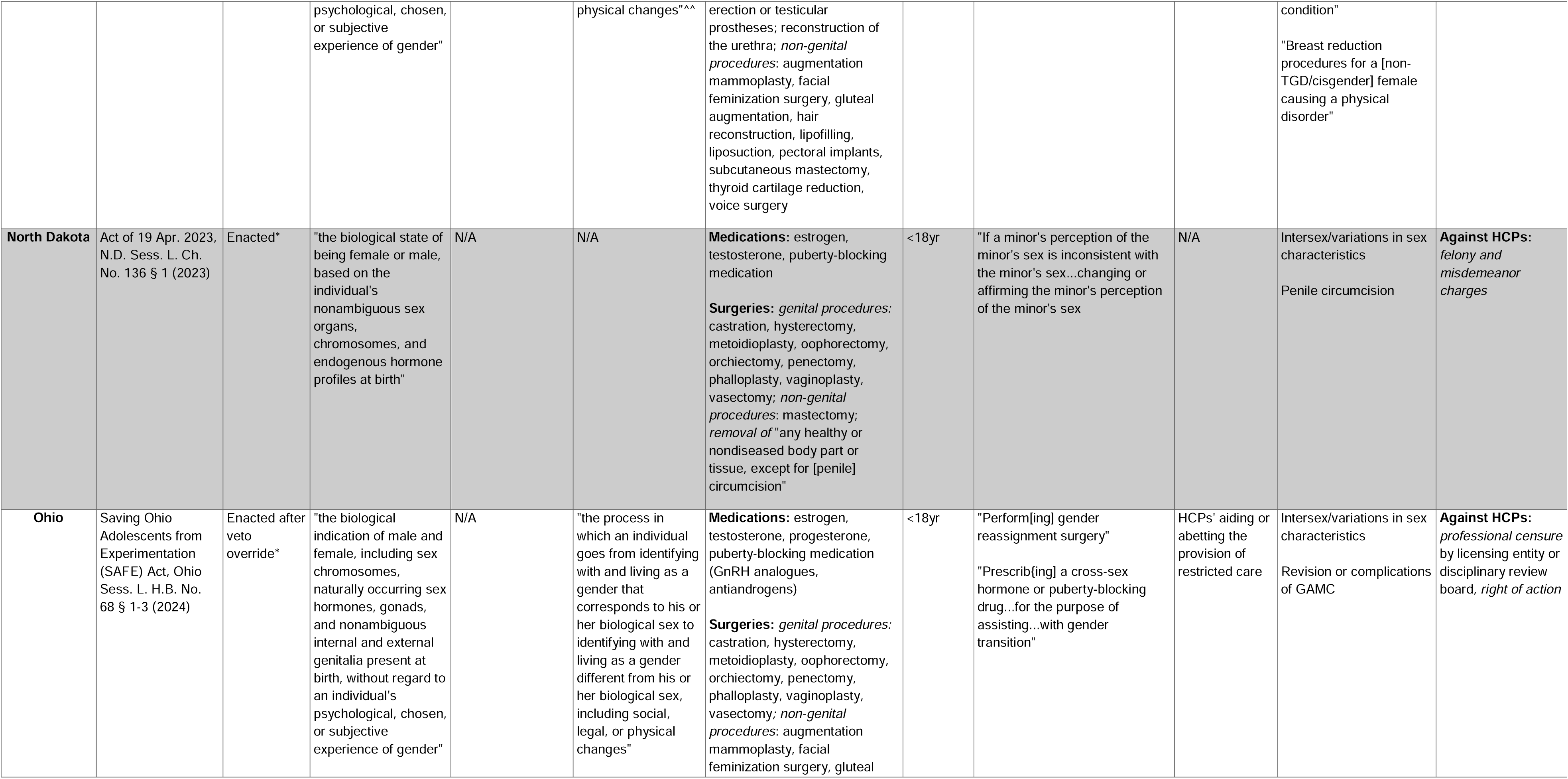
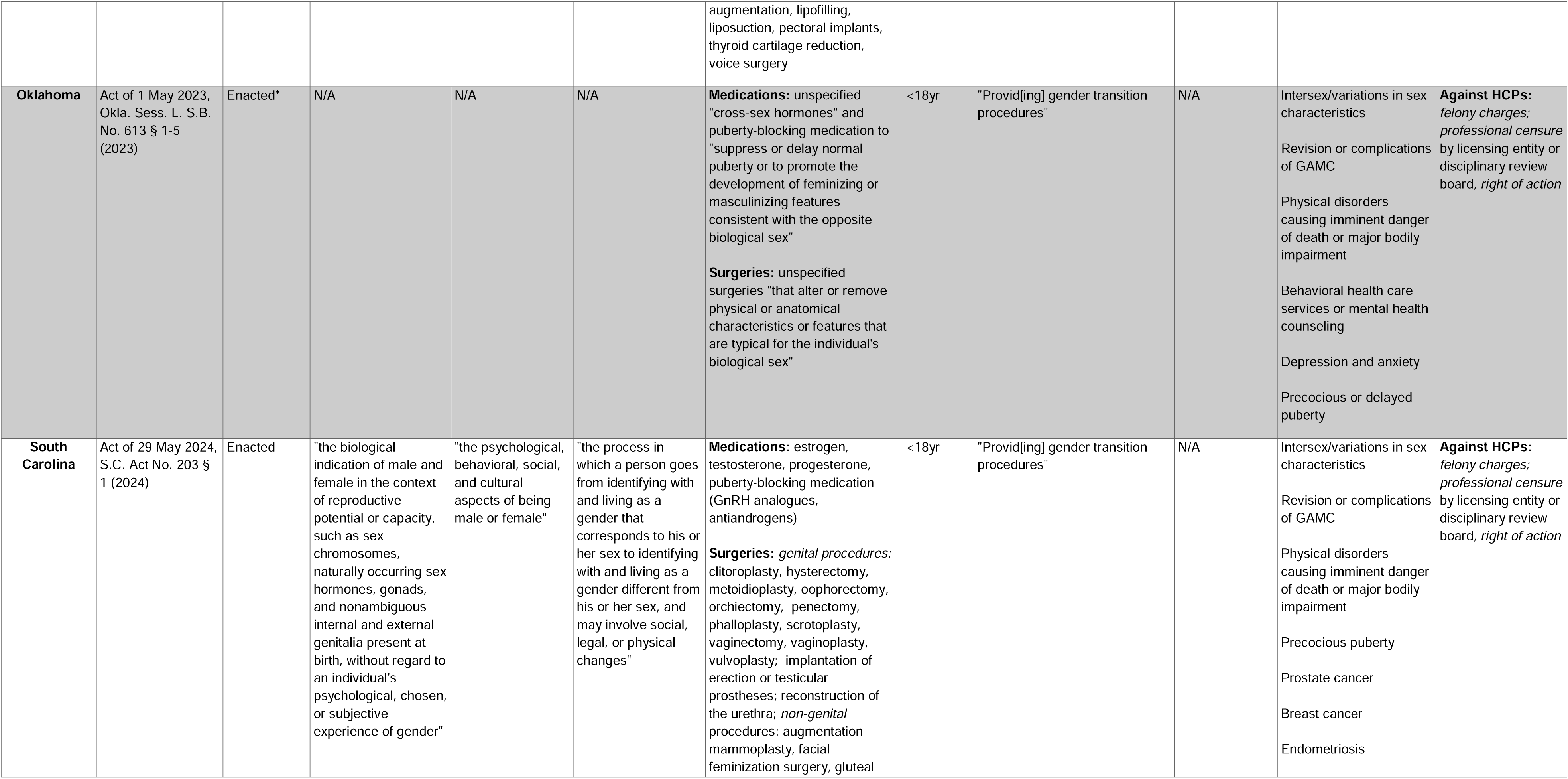
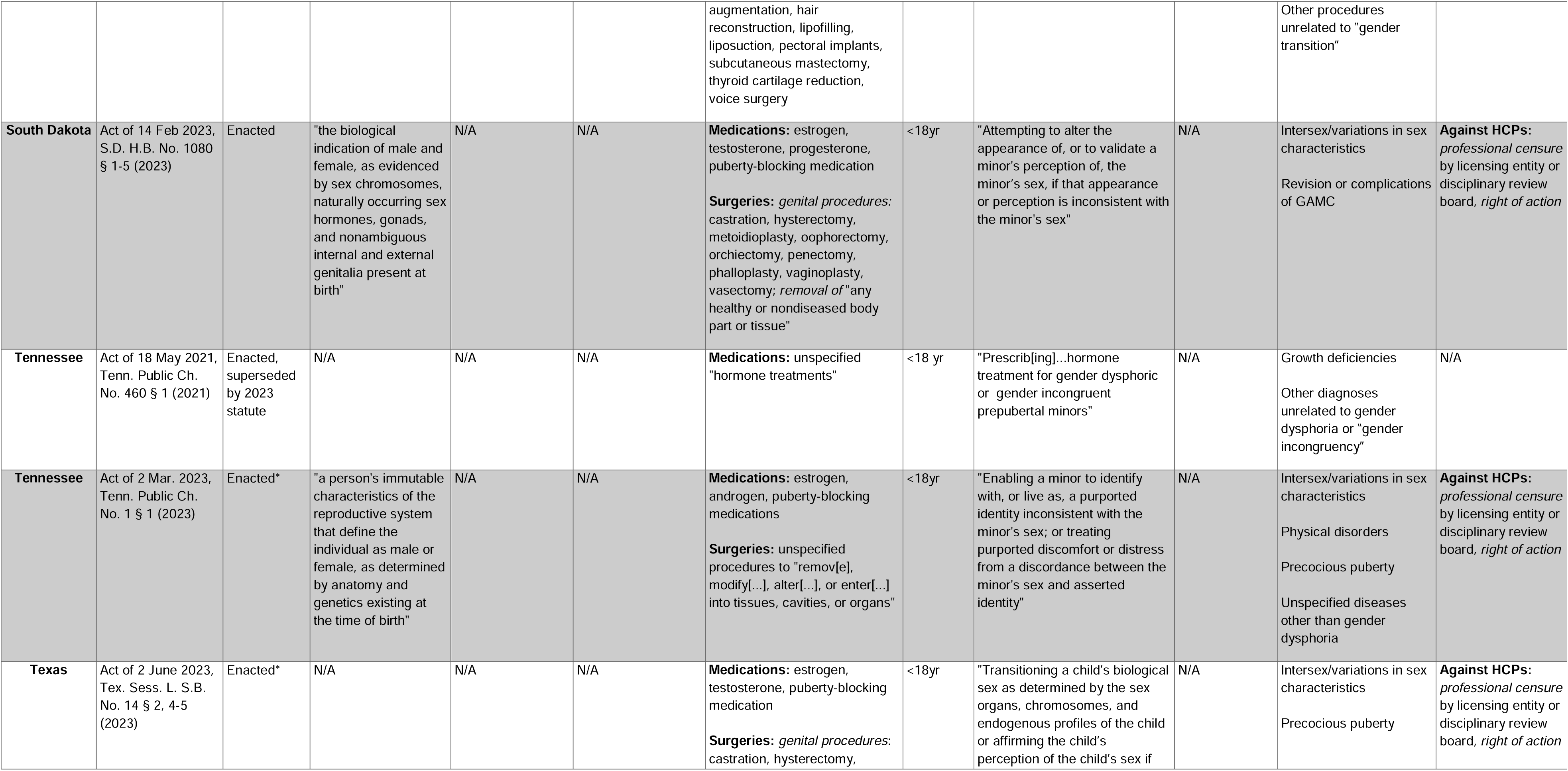
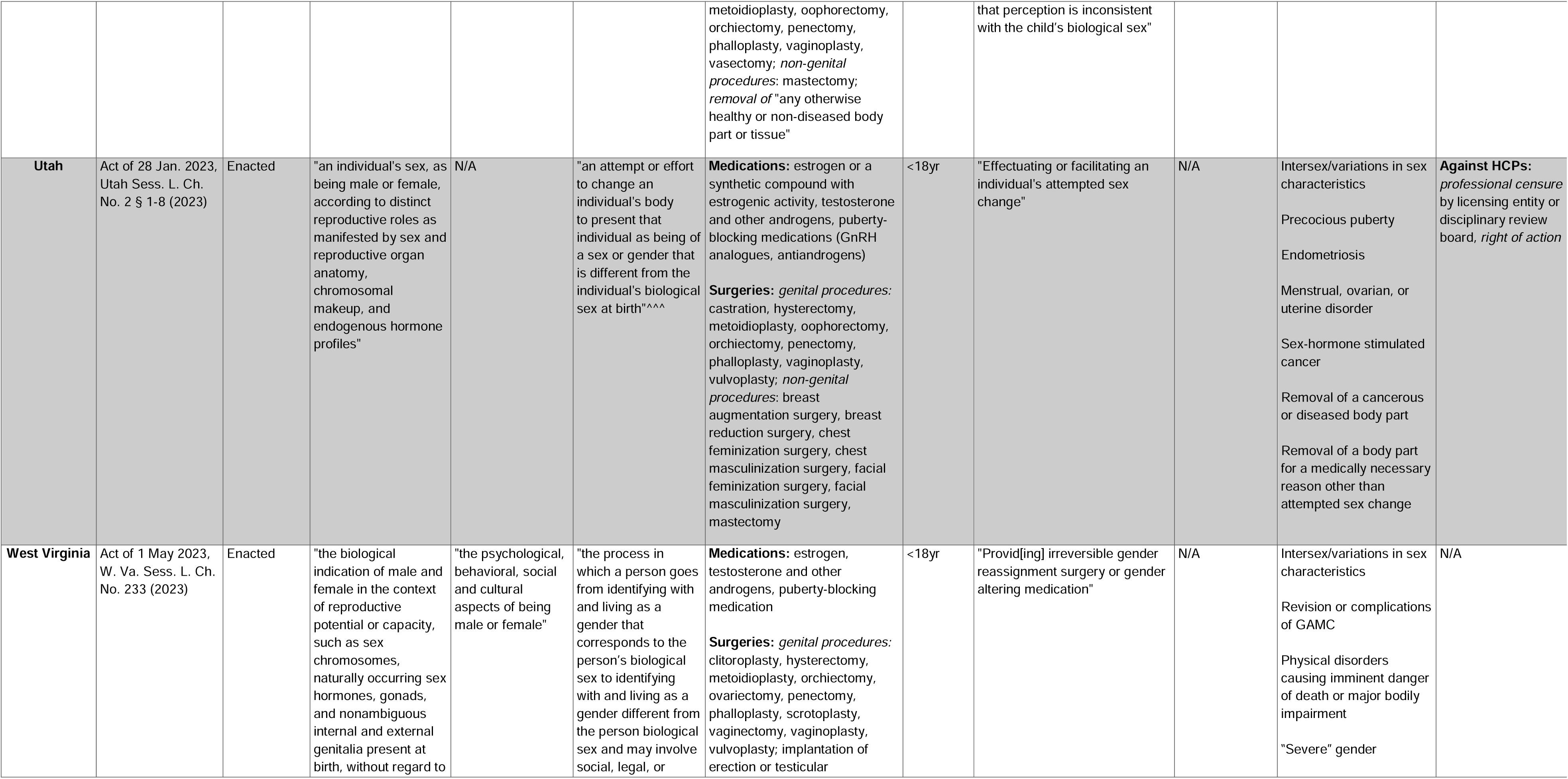
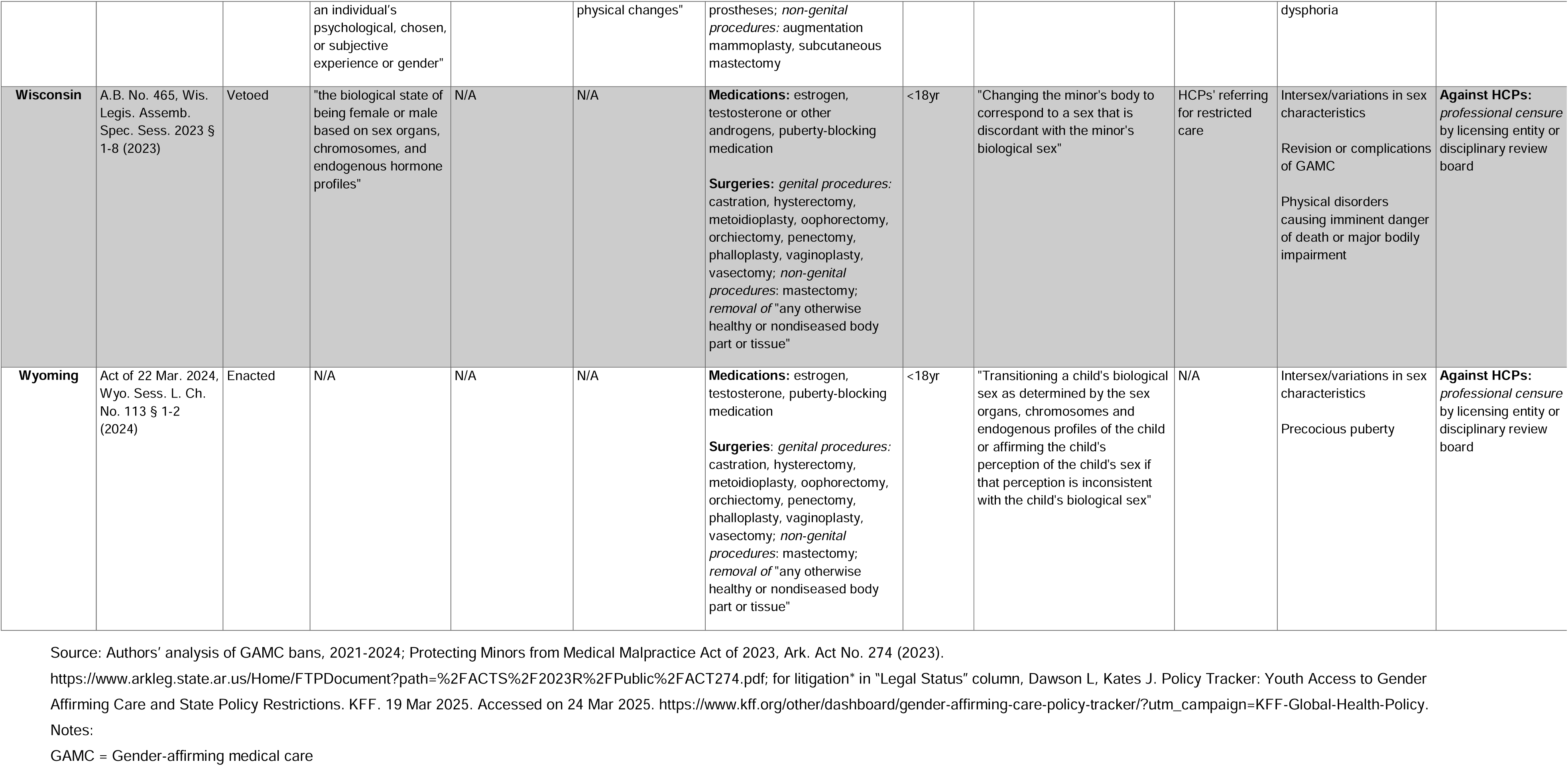
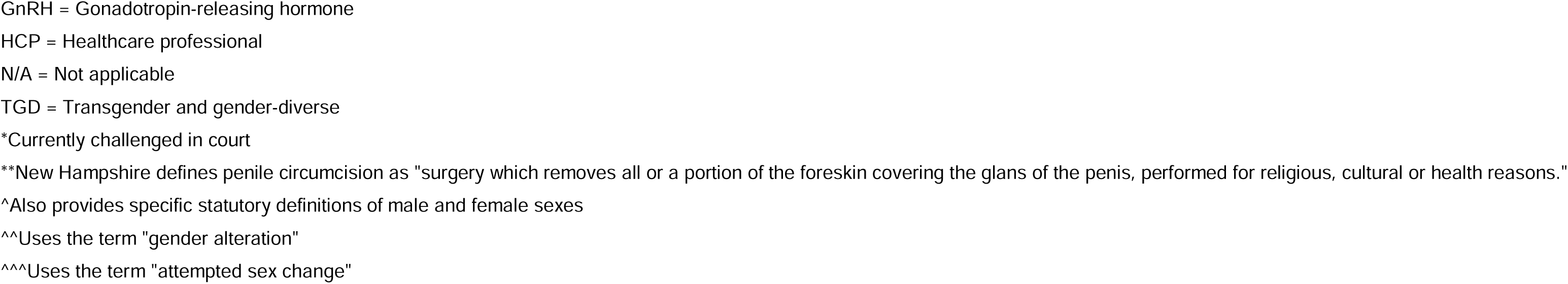
Elements of U.S. laws restricting medical care for transgender minors, 2021-2024 (N=30).

| State | Session Law (Legislative Year) | Legal Status | Statutory Definition of Sex | Statutory Definition of Gender | Statutory Definition of Gender Transition | Type of Medical Care Restricted | Ages Affected | Purpose(s) for Which Medical Care Is Restricted | Additional Restrictions | Statutory Exceptions | Penalties |
| --- | --- | --- | --- | --- | --- | --- | --- | --- | --- | --- | --- |
| Alabama | Alabama Vulnerable Child Compassion and Protection Act, Ala. Act No. 2022-289 § 1-4 (2022) | Enacted* | "the biological state of being female or male, based on the individual's sex organs, chromosomes, and endogenous hormone profiles" | N/A | N/A | **Medications:** estrogen, testosterone and other androgens, puberty-blocking medication<br>**Surgeries:** *genital procedures:* castration, hysterectomy, metoidioplasty, oophorectomy, orchiectomy, penectomy, phalloplasty, vaginoplasty, vasectomy; *removal of* "any healthy or non-diseased body part or tissue, except for a [penile] circumcision" | <19yr | "Attempting to alter the appearance of or affirm the minor's perception of his or her gender or sex, if that appearance or perception is inconsistent with the minor's sex as defined in this act" | N/A | Intersex/variations in sex characteristics<br>Penile circumcision | *Felony charges* for any person who "engage[s] in or cause[s] the [restricted medical] practices to be performed upon a minor" |
| Arizona | Act of 30 Mar. 2022, Ariz. Sess. L. Ch. No. 104 § 1 (2022) | Enacted | "the biological indication of male and female in the context of reproductive potential or capacity, such as sex chromosomes, naturally occurring sex hormones, gonads and nonambiguous internal and external genitalia present at birth, without regard to an individual's psychological, chosen or subjective experience of gender" | "the psychological, behavioral, social and cultural aspects of being male or female" | "the process in which a person goes from identifying with and living as a gender that corresponds to the person's biological sex to identifying with and living as a gender different from the person's biological sex and may involve social, legal or physical changes" | **Surgeries:** *genital procedures:* clitoroplasty, hysterectomy, metoidioplasty, penectomy, phalloplasty, orchiectomy, ovariectomy, scrotoplasty, vaginectomy, vaginoplasty, vulvoplasty; implantation of erection or testicular prostheses; *non-genital procedures:* augmentation mammoplasty, subcutaneous mastectomy | <18yr | "Assisting an individual with a gender transition" | N/A | Intersex/variations in sex characteristics<br>Revision or complications of GAMC<br>Physical disorders causing imminent danger of death or major bodily impairment | N/A |
| Arkansas | Arkansas Save Adolescents from Experimentation (SAFE) Act, Ark. | Enacted after veto override* | "the biological indication of male and female in the context of reproductive | "the psychological, behavioral, social, and cultural aspects of being | "the process in which a person goes from identifying with and living as a | **Medications:** estrogen, testosterone and other androgens, puberty-blocking medication (GnRH analogues) | <18yr | "Provid[ing] gender transition procedures" | HCPs' referring for restricted care | Intersex/variations in sex characteristics<br>Revision or complications | **Against HCPs:** *professional censure* by licensing entity or disciplinary review |
|  | Act No. 626 § 1-3 (2021) |  | potential or capacity, such as sex chromosomes, naturally occurring sex hormones, gonads, and nonambiguous internal and external genitalia present at birth, without regard to an individual's psychological, chosen, or subjective experience of gender" | male or female" | gender that corresponds to his or her biological sex to identifying with and living as a gender different from his or her biological sex and may involve social, legal or physical changes" | **Surgeries:** *genital procedures:* clitoroplasty, hysterectomy, metoidioplasty, orchiectomy, oophorectomy, penectomy, phalloplasty, scrotoplasty, vaginectomy, vaginoplasty, vulvoplasty; implantation of erection or testicular prostheses; reconstruction of the urethra; *non-genital procedures:* augmentation mammoplasty, facial feminization surgery, gluteal augmentation, hair reconstruction, lipofilling, liposuction, pectoral implants, subcutaneous mastectomy, thyroid cartilage reduction, voice surgery; *alteration or removal* of "biologically healthy and functional body parts" |  |  |  | of GAMC<br>Physical disorders causing imminent danger of death or major bodily impairment | board; *right of action*, including subsequent Protecting Minors from Medical Malpractice Act of 2023 (Ark. Act No. 274, 2023) |
| Florida | Act of 18 May 2023, Fla. Sess. L. Ch. No. 2023-90 § 4-7 (2023) | Enacted* | "the classification of a person as either male or female based on the organization of the human body of such person for a specific reproductive role, as indicated by the person's sex chromosomes, naturally occurring sex hormones, and internal and external genitalia present at birth" | N/A | N/A | **Medications:** unspecified hormones or hormone antagonists to affirm a person's perception of their sex if that perception is inconsistent with their statutorily defined sex; puberty-blocking medications<br>**Surgeries:** unspecified procedures to affirm a person's perception of their sex if that perception is inconsistent with their statutorily defined sex | <18yr | "Affirm[ing] a person's perception of his or her sex if that perception is inconsistent with the person's sex as defined in [the statute]" | N/A | Intersex/variations in sex characteristics<br>Revision or complications of GAMC<br>Physical disorders causing imminent danger of death or major bodily impairment | **Against HCPs:** *felony and misdemeanor charges, professional censure* by licensing entity or disciplinary review board, *right of action* against |
| Georgia | Act of 23 Mar. 2023, Ga. Act No. 4 § 1-3 (2023) | Enacted* | N/A | N/A | N/A | **Medications:** unspecified hormone replacement therapies<br>**Surgeries:** unspecified procedures that alter primary or | <18yr | "Treatment of gender dysphoria" | N/A | Intersex/variations in sex characteristics<br>Medically necessary conditions other than | **Against HCPs:** *professional censure* by licensing entity or disciplinary review |
|  |  |  |  |  |  | secondary sexual characteristics |  |  |  | gender dysphoria; or medically necessary sex reassignment<br>Partial androgen insensitivity syndrome | board |
| Idaho | Vulnerable Child Protection Act, Ida. Sess. L. Ch. No. 292 § 1-2 (2023) | Enacted* | "the immutable biological and physiological characteristics, specifically the chromosomes and internal and external reproductive anatomy, genetically determined at conception and generally recognizable at birth, that define an individual as male or female" | N/A | N/A | **Medications:** estrogen, testosterone, puberty-blocking medication<br>**Surgeries:** *genital surgeries:* castration, clitoroplasty, hysterectomy, metoidioplasty, oophorectomy, orchiectomy, ovariectomy, penectomy, phalloplasty, scrotoplasty, vaginoplasty, vasectomy, vulvoplasty; implantation of erection or testicular prostheses; reconstruction of the urethra; *non-genital procedures:* mastectomy; *removal of* "any otherwise healthy or nondiseased body part or tissue" | <18yr | "Attempting to alter the appearance of or affirm the child's perception of the child's sex if that perception is in consistent with the child's biological sex" | N/A | Intersex/variations in sex characteristics<br>Revision or complications of GAMC<br>Physical disorders causing imminent danger of death or major bodily impairment<br>Medical necessary conditions other than restricted circumstances | **Against HCPs:** *felony charges and fines* |
| Indiana | Act of 5 Apr. 2023, Ind. Pub. L. No. 10 (2023) | Enacted* | "the biological state of being male or female, based on the individual's sex organs, chromosomes, and endogenous hormone profiles" | "the psychological, behavioral, social, and cultural aspects of being male or female" | "the process in which an individual shifts from identifying with and living as a gender that corresponds to his or her sex to identifying with and living as a gender different from his or her sex, and may involve social, legal, or physical changes" | **Medications:** estrogen, testosterone, progesterone, puberty-blocking medication (GnRH analogues, antiandrogens)<br>**Surgeries:** *genital procedures:* clitoroplasty, hysterectomy, metoidioplasty, orchiectomy, ovariectomy, penectomy, phalloplasty, scrotoplasty, vaginectomy, vaginoplasty, vulvoplasty; implantation of erection or testicular prostheses; reconstruction of the urethra; *non-genital procedures:* augmentation mammoplasty, facial feminization surgery, gluteal augmentation, hair reconstruction, lipofilling, liposuction, pectoral implants, subcutaneous mastectomy, thyroid cartilage reduction, voice surgery | <18yr | "Providing gender transition procedures" | HCPs aiding or abetting the provision of restricted care | Intersex/variations in sex characteristics<br>Revision or complications of GAMC<br>Physical disorders causing imminent danger of death or major bodily impairment<br>Mental health or social services | **Against HCPs:** *professional censure by licensing entity or disciplinary review board, right of action* |
| **Iowa** | Act of 22 Mar. 2023, Iowa Sess. L. Ch. No. 9 § 1 (2023) | Enacted | "the biological indication of male and female, including sex chromosomes, naturally occurring sex hormones, gonads, and nonambiguous internal and external genitalia present at birth without regard to an individual's psychological, chosen, or subjective experience of gender" | "the psychological, behavioral, social, and cultural aspects of being male or female" | N/A | **Medications:** estrogen, testosterone, progesterone, puberty-blocking medication (GnRH analogues, antiandrogens)<br>**Surgeries:** *genital procedures:* castration, hysterectomy, metoidioplasty, oophorectomy, orchiectomy, penectomy, phalloplasty, vaginoplasty, vasectomy; *removal of* "any healthy or nondiseased body part or tissue" | <18yr | "Attempting to alter the appearance of, or affirm the minor's perception of, the minor's gender or sex, if that appearance or perception is inconsistent with the minor's sex" | HCPs' aiding or abetting the provision of restricted care | Intersex/variations in sex characteristics<br>Revision or complications of GAMC<br>Physical disorders causing imminent danger of death or major bodily impairment | **Against HCPs:** *professional censure* by licensing entity or disciplinary review board, *right of action* |
| **Kansas** | Kansas Child Mutilation Prevention Act, Kan. S.B. No. 26, Kan. Legis. Assemb. Reg. Sess. 2023 § 1-2 (2023) | Vetoed | "the biological state of being female or male based on the individual's sex organs, chromosomes and endogenous hormone profiles" | N/A | N/A | **Medications:** estrogen, testosterone, puberty-blocking medication<br>**Surgeries:** *genital procedures:* castration, hysterectomy, metoidioplasty, oophorectomy, orchiectomy, penectomy, phalloplasty, vaginoplasty, vasectomy; *non-genital procedures:* mastectomy; *removal of* "any body part or tissue" | <18yr | "Attempting to affirm the child's perception of the child's sex or gender, if that perception is inconsistent with the child's sex" | N/A | Intersex/variations in sex characteristics | **Against HCPs:** *professional censure* by licensing entity or disciplinary review board, *right of action* |
| **Kansas** | Forbidding Abusive Child Transitions Act, Kan. S.B. 233, Kan. Legis. Assemb. Reg. Sess. 2024 § 1, 3-6 | Vetoed | "the biological indication of male and female in the context of reproductive potential or capacity, including sex | "the psychological, behavioral, social and cultural aspects of being male or female" | N/A | **Medications:** estrogen, testosterone and other androgens, puberty-blocking medication (GnRH analogues)<br>**Surgeries:** *genital procedures:* | <18yr | "Provid[ing] the [restricted care] to a female child whose perceived gender or sex is not female as treatment for distress arising from such female child's perception that such child's | N/A | Intersex/variations in sex characteristics<br>Revision or complications of GAMC | **Against HCPs:** *professional censure* by licensing entity or disciplinary review board, *right of action*, *prohibition on* |
|  | (2024) |  | chromosomes, naturally occurring sex hormones, gonads and nonambiguous internal and external genitalia present at birth, without regard to an individual's psychological, chosen or subjective experience of gender" |  |  | clitoroplasty, hysterectomy, metoidioplasty, oophorectomy, orchiectomy, ovariectomy, penectomy, phalloplasty, scrotoplasty, vaginectomy, vaginoplasty, vulvoplasty; implantation of erection or testicular prostheses; reconstruction of the urethra; *non-genital procedures*: augmentation mammoplasty, facial feminization surgery, gluteal augmentation, lipofilling, liposuction, pectoral implants, subcutaneous mastectomy, thyroid cartilage reduction, voice surgery |  | gender or sex is not female"<br>"Provid[ing] the [restricted care] to a male child whose perceived gender or sex is not male as treatment for distress arising from such male child's perception that such child's gender or sex is not male" |  |  | *professional liability insurance coverage* for restricted care |
| **Kentucky** | Act of 29 Mar. 2023, Ky. Sess. L. Ch. No. 132 § 4 (2023) | Enacted after veto override* | "the biological indication of male and female as evidenced by sex chromosomes, naturally occurring sex hormones, gonads, and nonambiguous internal and external genitalia present at birth" | N/A | N/A | **Medications:** estrogen, testosterone, progesterone, puberty-blocking medication<br>**Surgeries:** *genital procedures*: castration, hysterectomy, metoidioplasty, oophorectomy, orchiectomy, penectomy, phalloplasty, vaginoplasty, vasectomy, *removal of* "any healthy or non-diseased body part or tissue" | <18yr | "Attempting to alter the appearance of, or to validate a minor's perception of, the minor's sex, if that appearance or perception is inconsistent with the minor's sex" | N/A | Intersex/variations in sex characteristics<br>Revision or complications of GAMC | **Against HCPs:** *professional censure* by licensing entity or disciplinary review board, *right of action* |
| **Louisiana** | The Stop Harming Our Kids Act, La. Act No. 466 § 1 (2023) | Enacted after veto override* | "the biological indication of male or female as evidenced by a medical examination of all of the following:<br>(a) Sex chromosomes<br>(b) Naturally occurring sex hormones<br>(c) Gonads<br>(d) As well as nonambiguous internal and external genitalia present at birth" | N/A | N/A | **Medications:** estrogen, testosterone, progesterone, puberty-blocking medication (GnRH analogues, antiandrogens)<br>**Surgeries:** *genital procedures*: castration, hysterectomy, metoidioplasty, oophorectomy, orchiectomy, penectomy, phalloplasty, vaginoplasty, vasectomy; *non-genital procedures*: augmentation mammoplasty, facial feminization surgery, gluteal augmentation, hair reconstruction, lipofilling, liposuction, pectoral implants, thyroid cartilage reduction, voice surgery; *removal of* "any healthy or non-diseased body part or tissue" | <18yr | "Attempt[ing] to alter a minor's appearance in an attempt to validate a minor's perception of the minor's sex, if the minor's perception is inconsistent with the minor's sex" | N/A | Intersex/variations in sex characteristics<br>Revision or complications of GAMC<br>Physical disorders causing imminent danger of death or major bodily impairment | **Against HCPs:** *professional censure* by licensing entity or disciplinary review board, *right of action* |
| **Mississippi** | Regulate Experimental Adolescent Procedures (REAP) Act, Miss. Sess. L. Ch. No. 303 § 1-3, 5 (2023) | Enacted | "the biological indication of male and female in the context of reproductive potential or capacity, such as sex chromosomes, naturally occurring sex hormones, gonads, and nonambiguous internal and external genitalia present at birth, without regard to an individual's psychological, chosen, or subjective experience of gender" | "the psychological, behavioral, social, and cultural aspects of being male or female" | "the process in which a person goes from identifying with and living as a gender that corresponds to his or her sex to identifying with and living as a gender different from his or her sex, and may involve social, legal, or physical changes" | **Medications:** estrogen, testosterone and other androgens, puberty-blocking medication (GnRH analogues)<br>**Surgeries:** *genital surgeries:* castration, clitoroplasty, hysterectomy, metoidioplasty, oophorectomy, orchiectomy, penectomy, phalloplasty, scrotoplasty, vaginectomy, vaginoplasty, vulvoplasty; implantation of erection or testicular prostheses; reconstruction of the urethra; *non-genital* procedures: augmentation mammoplasty, facial feminization surgery, gluteal augmentation, hair reconstruction, lipofilling, liposuction, pectoral implants, subcutaneous mastectomy, thyroid cartilage reduction, voice surgery | <18yr | "Provid[ing] gender transition procedures" | HCPs' aiding or abetting the provision of restricted care | Intersex/variations in sex characteristics<br>Revision or complications of GAMC<br>Penile circumcision | **Against HCPs:** *professional censure* by licensing entity or disciplinary review board, *right of action* |
| **Missouri** | Missouri Save Adolescents from Experimentation (SAFE) Act, Mo. S. Sub. No. 2 § A (2023) | Enacted* | "the biological indication of male or female in the context of reproductive potential or capacity, such as sex chromosomes, naturally occurring sex hormones, gonads, | "the psychological, behavioral, social, and cultural aspects of being male or female" | "the process in which an individual transitions from identifying with and living as a gender that corresponds to his or her biological sex to identifying with and living as a | **Medications:** estrogen, testosterone and other androgens, puberty-blocking medications (GnRH analogues, antiandrogens)<br>**Surgeries:** *genital procedures:* castration, hysterectomy, metoidioplasty, oophorectomy, | <18yr | "[Providing restricted care for] a gender transition" | N/A | Intersex/variations in sex characteristics<br>Revision or complications of GAMC<br>Physical disorders causing imminent danger of death or major bodily | **Against HCPs:** *professional censure* by licensing entity or disciplinary review board, *right of action* |
|  |  |  | and nonambiguous internal and external genitalia present at birth, without regard to an individual's psychological, chosen, or subjective experience of gender" |  | gender different from his or her biological sex, and may involve social, legal, or physical changes" | orchiectomy, penectomy, phalloplasty, vaginoplasty, vasectomy; *non-genital procedures*: augmentation mammoplasty, subcutaneous mastectomy |  |  |  | impairment |  |
| Montana | Youth Health Protection Act, Mont. Sess. L. Ch. No. 306 § 1-7 (2023) | Enacted* | "the organization of body parts and gametes for reproduction in human beings and other organisms. In human beings, there are exactly two sexes, male and female, with two corresponding types of gametes. The sexes are determined by the biological and genetic indication of male or female, including sex chromosomes, naturally occurring sex chromosomes, gonads, and nonambiguous internal and external genitalia present at birth, without regard to an individual's psychological, behavioral, social, cultural, chosen, or subjective experience of gender" | "the psychological, behavioral, social, and cultural aspects of being male or female. An individual's gender may or may not align with the individual's sex"^ | N/A | **Medications:** estrogen, testosterone and other androgens, puberty-blocking medication (GnRH analogues)<br>**Surgeries:** *genital procedures*: clitoroplasty, hysterectomy, metoidioplasty, oophorectomy, orchiectomy, ovariectomy, penectomy, phalloplasty, scrotoplasty, vaginectomy, vaginoplasty, vulvoplasty; implantation of erection or testicular prostheses; reconstruction of the urethra; *non-genital procedures*: augmentation mammoplasty, facial feminization surgery, gluteal augmentation, pectoral implants, subcutaneous mastectomy, thyroid cartilage reduction, voice surgery | <18yr | "Provid[ing] [the restricted care] to address a female minor's perception that her gender or sex is not female or a male minor's perception that his gender or sex is not male" | N/A | Intersex/variations in sex characteristics<br>Revision or complications of GAMC | **Against HCPs:** *professional censure* by licensing entity or disciplinary review board, *right of action*, *prohibition on professional liability insurance coverage* for restricted care |
| Nebraska | Let Them Grow Act, Neb. Sess. L. L.B. No. 574 § 14-18, 20 | Enacted* | "the biological indication of male and female in the context of reproductive | "the psychological, behavioral, social, and cultural aspects of being | "the process in which a person goes from identifying with and living as a | **Medications:** estrogen, testosterone and other androgens, puberty-blocking medication (GnRH analogues) | <19yr | "Perform[ing] gender-altering procedures" | N/A | Intersex/variations in sex characteristics<br>Revision or complications | **Against HCPs:** *professional censure* by licensing entity or disciplinary review |
|  | (2023) |  | potential or capacity, such as sex chromosomes, naturally occurring sex hormones, gonads, and nonambiguous internal and external genitalia present at birth, without regard to an individual's psychological, chosen, or subjective experience of gender" | male or female" | gender that corresponds to his or her biological sex to identifying with and living as a gender different from his or her biological sex and may involve social, legal, or physical changes"^^ | **Surgeries:** *genital procedures:* clitoroplasty, hysterectomy, metoidioplasty, orchiectomy, ovariectomy, penectomy, phalloplasty, scrotoplasty, vaginectomy, vaginoplasty, vulvoplasty; implantation of erection or testicular prostheses; reconstruction of the urethra; *non-genital procedures:* subcutaneous mastectomy, thyroid cartilage reduction, voice surgery |  |  |  | of GAMC<br>Physical disorders causing imminent danger of death or major bodily impairment | board, *right of action* |
| New Hampshire | Act of 26 July 2024, N.H. Sess. L. Ch. No. 213 (2024) | Enacted | N/A | N/A | N/A | **Surgeries:** *genital procedures:* metoidioplasty, phalloplasty, or vaginoplasty | <18yr | "Perform[ing] genital gender reassignment surgery" | HCPs' referring for restricted care | Intersex/variations in sex characteristics<br>Penile circumcision**<br>Genital reconstructive surgeries to "correct malformation, malignancy, injury or physical disease"<br>"Removal of malignant, malformed, or otherwise damaged genitalia" | **Against HCPs:** *professional censure* by licensing entity or disciplinary review board, *right of action* |
| North Carolina | Act of 16 Aug. 2023, N.C. Sess. L. Ch. No. 2023-111 § 1-2 (2023) | Enacted after veto override* | "the biological indication of male and female in the context of reproductive potential or capacity, such as sex chromosomes, naturally occurring sex hormones, gonads, and nonambiguous internal and external genitalia present at birth, without regard to an individual's | "the psychological, behavioral, social, and cultural aspects of being male or female" | "the process in which a person goes from identifying with and living as a gender that corresponds to his or her biological sex to identifying with and living as a gender different from his or her biological sex and may involve social, legal, or | **Medications:** estrogen or synthetic compounds with estrogenic activity, testosterone and other androgens, puberty-blocking medication (GnRH analogues)<br>**Surgeries:** *genital procedures:* clitoroplasty, hysterectomy, metoidioplasty, orchiectomy, ovariectomy, penectomy, phalloplasty, scrotoplasty, vaginectomy, vaginoplasty, vulvoplasty; implantation of | <18yr | "Perform[ing] a surgical gender transition procedure...[and] prescrib[ing], provid[ing] or dispens[ing] puberty-blocking drugs or cross-sex hormones" | N/A | Intersex/variations in sex characteristics<br>Revision or complications of GAMC<br>Physical disorders causing imminent danger of death or major bodily impairment<br>"Any surgery...[that] is medically necessary to treat a physiological | **Against HCPs:** *professional censure* by licensing entity or disciplinary review board, *right of action* |
|  |  |  | psychological, chosen, or subjective experience of gender" |  | physical changes"^^ | erection or testicular prostheses; reconstruction of the urethra; *non-genital procedures*: augmentation mammoplasty, facial feminization surgery, gluteal augmentation, hair reconstruction, lipofilling, liposuction, pectoral implants, subcutaneous mastectomy, thyroid cartilage reduction, voice surgery |  |  |  | condition" |  |
| North Dakota | Act of 19 Apr. 2023, N.D. Sess. L. Ch. No. 136 § 1 (2023) | Enacted* | "the biological state of being female or male, based on the individual's nonambiguous sex organs, chromosomes, and endogenous hormone profiles at birth" | N/A | N/A | **Medications:** estrogen, testosterone, puberty-blocking medication<br>**Surgeries:** *genital procedures*: castration, hysterectomy, metoidioplasty, oophorectomy, orchiectomy, penectomy, phalloplasty, vaginoplasty, vasectomy; *non-genital procedures*: mastectomy; *removal of* "any healthy or nondiseased body part or tissue, except for [penile] circumcision" | <18yr | "If a minor's perception of the minor's sex is inconsistent with the minor's sex...changing or affirming the minor's perception of the minor's sex | N/A | Intersex/variations in sex characteristics<br>Penile circumcision | **Against HCPs:** *felony and misdemeanor charges* |
| Ohio | Saving Ohio Adolescents from Experimentation (SAFE) Act, Ohio Sess. L. H.B. No. 68 § 1-3 (2024) | Enacted after veto override* | "the biological indication of male and female, including sex chromosomes, naturally occurring sex hormones, gonads, and nonambiguous internal and external genitalia present at birth, without regard to an individual's psychological, chosen, or subjective experience of gender" | N/A | "the process in which an individual goes from identifying with and living as a gender that corresponds to his or her biological sex to identifying with and living as a gender different from his or her biological sex, including social, legal, or physical changes" | **Medications:** estrogen, testosterone, progesterone, puberty-blocking medication (GnRH analogues, antiandrogens)<br>**Surgeries:** *genital procedures*: castration, hysterectomy, metoidioplasty, oophorectomy, orchiectomy, penectomy, phalloplasty, vaginoplasty, vasectomy; *non-genital procedures*: augmentation mammoplasty, facial feminization surgery, gluteal augmentation, lipofilling, liposuction, pectoral implants, thyroid cartilage reduction, voice surgery | <18yr | "Perform[ing] gender reassignment surgery"<br>"Prescrib[ing] a cross-sex hormone or puberty-blocking drug...for the purpose of assisting...with gender transition" | HCPs' aiding or abetting the provision of restricted care | Intersex/variations in sex characteristics<br>Revision or complications of GAMC | **Against HCPs:** *professional censure* by licensing entity or disciplinary review board, *right of action* |
| Oklahoma | Act of 1 May 2023, Okla. Sess. L. S.B. No. 613 § 1-5 (2023) | Enacted* | N/A | N/A | N/A | **Medications:** unspecified "cross-sex hormones" and puberty-blocking medication to "suppress or delay normal puberty or to promote the development of feminizing or masculinizing features consistent with the opposite biological sex"<br>**Surgeries:** unspecified surgeries "that alter or remove physical or anatomical characteristics or features that are typical for the individual's biological sex" | <18yr | "Provid[ing] gender transition procedures" | N/A | Intersex/variations in sex characteristics<br>Revision or complications of GAMC<br>Physical disorders causing imminent danger of death or major bodily impairment<br>Behavioral health care services or mental health counseling<br>Depression and anxiety<br>Precocious or delayed puberty | **Against HCPs:**<br>*felony charges; professional censure* by licensing entity or disciplinary review board, *right of action* |
| South Carolina | Act of 29 May 2024, S.C. Act No. 203 § 1 (2024) | Enacted | "the biological indication of male and female in the context of reproductive potential or capacity, such as sex chromosomes, naturally occurring sex hormones, gonads, and nonambiguous internal and external genitalia present at birth, without regard to an individual's psychological, chosen, or subjective experience of gender" | "the psychological, behavioral, social, and cultural aspects of being male or female" | "the process in which a person goes from identifying with and living as a gender that corresponds to his or her sex to identifying with and living as a gender different from his or her sex, and may involve social, legal, or physical changes" | **Medications:** estrogen, testosterone, progesterone, puberty-blocking medication (GnRH analogues, antiandrogens)<br>**Surgeries:** *genital procedures:* clitoroplasty, hysterectomy, metoidioplasty, oophorectomy, orchiectomy, penectomy, phalloplasty, scrotoplasty, vaginectomy, vaginoplasty, vulvoplasty; implantation of erection or testicular prostheses; reconstruction of the urethra; *non-genital* procedures: augmentation mammoplasty, facial feminization surgery, gluteal | <18yr | "Provid[ing] gender transition procedures" | N/A | Intersex/variations in sex characteristics<br>Revision or complications of GAMC<br>Physical disorders causing imminent danger of death or major bodily impairment<br>Precocious puberty<br>Prostate cancer<br>Breast cancer<br>Endometriosis | **Against HCPs:**<br>*felony charges; professional censure* by licensing entity or disciplinary review board, *right of action* |
|  |  |  |  |  |  | augmentation, hair reconstruction, lipofilling, liposuction, pectoral implants, subcutaneous mastectomy, thyroid cartilage reduction, voice surgery |  |  |  | Other procedures unrelated to "gender transition" |  |
| **South Dakota** | Act of 14 Feb 2023, S.D. H.B. No. 1080 § 1-5 (2023) | Enacted | "the biological indication of male and female, as evidenced by sex chromosomes, naturally occurring sex hormones, gonads, and nonambiguous internal and external genitalia present at birth" | N/A | N/A | **Medications:** estrogen, testosterone, progesterone, puberty-blocking medication<br>**Surgeries:** *genital procedures:* castration, hysterectomy, metoidioplasty, oophorectomy, orchiectomy, penectomy, phalloplasty, vaginoplasty, vasectomy; *removal of* "any healthy or nondiseased body part or tissue" | <18yr | "Attempting to alter the appearance of, or to validate a minor's perception of, the minor's sex, if that appearance or perception is inconsistent with the minor's sex" | N/A | Intersex/variations in sex characteristics<br>Revision or complications of GAMC | **Against HCPs:** *professional censure* by licensing entity or disciplinary review board, *right of action* |
| **Tennessee** | Act of 18 May 2021, Tenn. Public Ch. No. 460 § 1 (2021) | Enacted, superseded by 2023 statute | N/A | N/A | N/A | **Medications:** unspecified "hormone treatments" | <18 yr | "Prescrib[ing]...hormone treatment for gender dysphoric or gender incongruent prepubertal minors" | N/A | Growth deficiencies<br>Other diagnoses unrelated to gender dysphoria or "gender incongruency" | N/A |
| **Tennessee** | Act of 2 Mar. 2023, Tenn. Public Ch. No. 1 § 1 (2023) | Enacted* | "a person's immutable characteristics of the reproductive system that define the individual as male or female, as determined by anatomy and genetics existing at the time of birth" | N/A | N/A | **Medications:** estrogen, androgen, puberty-blocking medications<br>**Surgeries:** unspecified procedures to "remov[e], modify[...], alter[...], or enter[...]" into tissues, cavities, or organs" | <18yr | "Enabling a minor to identify with, or live as, a purported identity inconsistent with the minor's sex; or treating purported discomfort or distress from a discordance between the minor's sex and asserted identity" | N/A | Intersex/variations in sex characteristics<br>Physical disorders<br>Precocious puberty<br>Unspecified diseases other than gender dysphoria | **Against HCPs:** *professional censure* by licensing entity or disciplinary review board, *right of action* |
| **Texas** | Act of 2 June 2023, Tex. Sess. L. S.B. No. 14 § 2, 4-5 (2023) | Enacted* | N/A | N/A | N/A | **Medications:** estrogen, testosterone, puberty-blocking medication<br>**Surgeries:** *genital procedures:* castration, hysterectomy, metoidioplasty, oophorectomy, orchiectomy, penectomy, phalloplasty, vaginoplasty, vasectomy; *non-genital procedures*: mastectomy; *removal of* "any otherwise healthy or non-diseased body part or tissue" | <18yr | "Transitioning a child's biological sex as determined by the sex organs, chromosomes, and endogenous profiles of the child or affirming the child's perception of the child's sex if that perception is inconsistent with the child's biological sex" | N/A | Intersex/variations in sex characteristics<br>Precocious puberty | **Against HCPs:** *professional censure* by licensing entity or disciplinary review board, *right of action* |
| **Utah** | Act of 28 Jan. 2023, Utah Sess. L. Ch. No. 2 § 1-8 (2023) | Enacted | "an individual's sex, as being male or female, according to distinct reproductive roles as manifested by sex and reproductive organ anatomy, chromosomal makeup, and endogenous hormone profiles" | N/A | "an attempt or effort to change an individual's body to present that individual as being of a sex or gender that is different from the individual's biological sex at birth"^^ | **Medications:** estrogen or a synthetic compound with estrogenic activity, testosterone and other androgens, puberty-blocking medications (GnRH analogues, antiandrogens)<br>**Surgeries:** *genital procedures*: castration, hysterectomy, metoidioplasty, oophorectomy, orchiectomy, penectomy, phalloplasty, vaginoplasty, vulvoplasty; *non-genital procedures*: breast augmentation surgery, breast reduction surgery, chest feminization surgery, chest masculinization surgery, facial feminization surgery, facial masculinization surgery, mastectomy | <18yr | "Effectuating or facilitating an individual's attempted sex change" | N/A | Intersex/variations in sex characteristics<br>Precocious puberty<br>Endometriosis<br>Menstrual, ovarian, or uterine disorder<br>Sex-hormone stimulated cancer<br>Removal of a cancerous or diseased body part<br>Removal of a body part for a medically necessary reason other than attempted sex change | **Against HCPs:** *professional censure* by licensing entity or disciplinary review board, *right of action* |
| **West Virginia** | Act of 1 May 2023, W. Va. Sess. L. Ch. No. 233 (2023) | Enacted | "the biological indication of male and female in the context of reproductive potential or capacity, such as sex chromosomes, naturally occurring sex hormones, gonads, and nonambiguous internal and external genitalia present at birth, without regard to | "the psychological, behavioral, social and cultural aspects of being male or female" | "the process in which a person goes from identifying with and living as a gender that corresponds to the person's biological sex to identifying with and living as a gender different from the person biological sex and may involve social, legal, or | **Medications:** estrogen, testosterone and other androgens, puberty-blocking medication<br>**Surgeries:** *genital procedures*: clitoroplasty, hysterectomy, metoidioplasty, orchiectomy, ovariectomy, penectomy, phalloplasty, scrotoplasty, vaginectomy, vaginoplasty, vulvoplasty; implantation of erection or testicular | <18yr | "Provid[ing] irreversible gender reassignment surgery or gender altering medication" | N/A | Intersex/variations in sex characteristics<br>Revision or complications of GAMC<br>Physical disorders causing imminent danger of death or major bodily impairment<br>"Severe" gender | N/A |
|  |  |  | an individual's psychological, chosen, or subjective experience or gender" |  | physical changes" | prostheses; *non-genital procedures*: augmentation mammoplasty, subcutaneous mastectomy |  |  |  | dysphoria |  |
| Wisconsin | A.B. No. 465, Wis. Legis. Assemb. Spec. Sess. 2023 § 1-8 (2023) | Vetoed | "the biological state of being female or male based on sex organs, chromosomes, and endogenous hormone profiles" | N/A | N/A | **Medications:** estrogen, testosterone or other androgens, puberty-blocking medication<br>**Surgeries:** *genital procedures*: castration, hysterectomy, metoidioplasty, oophorectomy, orchiectomy, penectomy, phalloplasty, vaginoplasty, vasectomy; *non-genital procedures*: mastectomy; *removal of* "any otherwise healthy or nondiseased body part or tissue" | <18yr | "Changing the minor's body to correspond to a sex that is discordant with the minor's biological sex" | HCPs' referring for restricted care | Intersex/variations in sex characteristics<br>Revision or complications of GAMC<br>Physical disorders causing imminent danger of death or major bodily impairment | **Against HCPs:** *professional censure* by licensing entity or disciplinary review board |
| Wyoming | Act of 22 Mar. 2024, Wyo. Sess. L. Ch. No. 113 § 1-2 (2024) | Enacted | N/A | N/A | N/A | **Medications:** estrogen, testosterone, puberty-blocking medication<br>**Surgeries:** *genital procedures*: castration, hysterectomy, metoidioplasty, oophorectomy, orchiectomy, penectomy, phalloplasty, vaginoplasty, vasectomy; *non-genital procedures*: mastectomy; *removal of* "any otherwise healthy or nondiseased body part or tissue" | <18yr | "Transitioning a child's biological sex as determined by the sex organs, chromosomes and endogenous profiles of the child or affirming the child's perception of the child's sex if that perception is inconsistent with the child's biological sex" | N/A | Intersex/variations in sex characteristics<br>Precocious puberty | **Against HCPs:** *professional censure* by licensing entity or disciplinary review board |

Source: Authors' analysis of GAMC bans, 2021-2024; Protecting Minors from Medical Malpractice Act of 2023, Ark. Act No. 274 (2023). https://www.arkleg.state.ar.us/Home/FTPDocument?path=%2FACTS%2F2023R%2FPublic%2FACT274.pdf; for litigation\* in "Legal Status" column, Dawson L, Kates J. Policy Tracker: Youth Access to Gender Affirming Care and State Policy Restrictions. KFF. 19 Mar 2025. Accessed on 24 Mar 2025. https://www.kff.org/other/dashboard/gender-affirming-care-policy-tracker/?utm_campaign=KFF-Global-Health-Policy. Notes: GAMC = Gender-affirming medical care
GnRH = Gonadotropin-releasing hormone
HCP = Healthcare professional
N/A = Not applicable
TGD = Transgender and gender-diverse
\*Currently challenged in court
\*\*New Hampshire defines penile circumcision as "surgery which removes all or a portion of the foreskin covering the glans of the penis, performed for religious, cultural or health reasons."
^Also provides specific statutory definitions of male and female sexes
^^Uses the term "gender alteration"

**Appendix Table 3.**
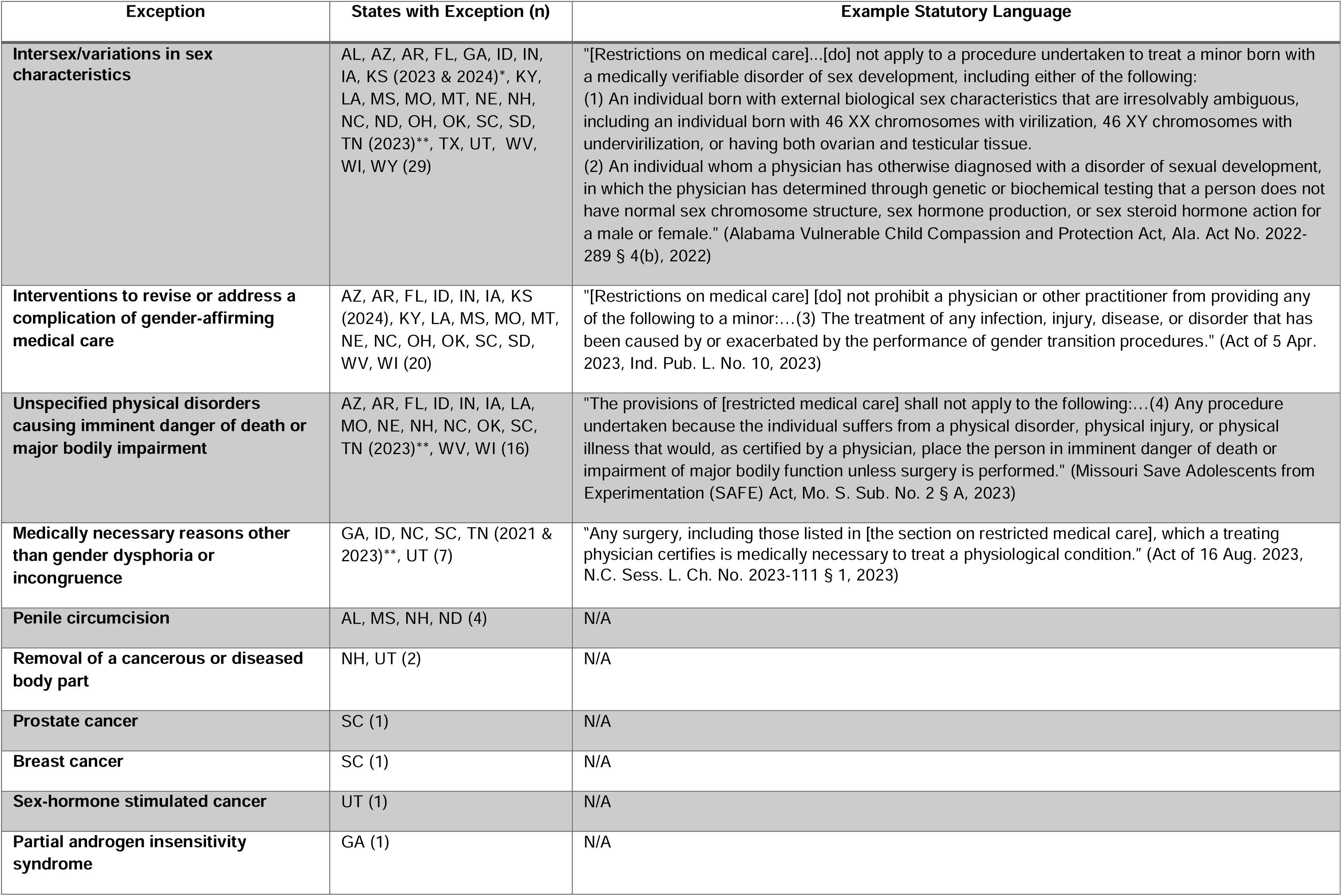
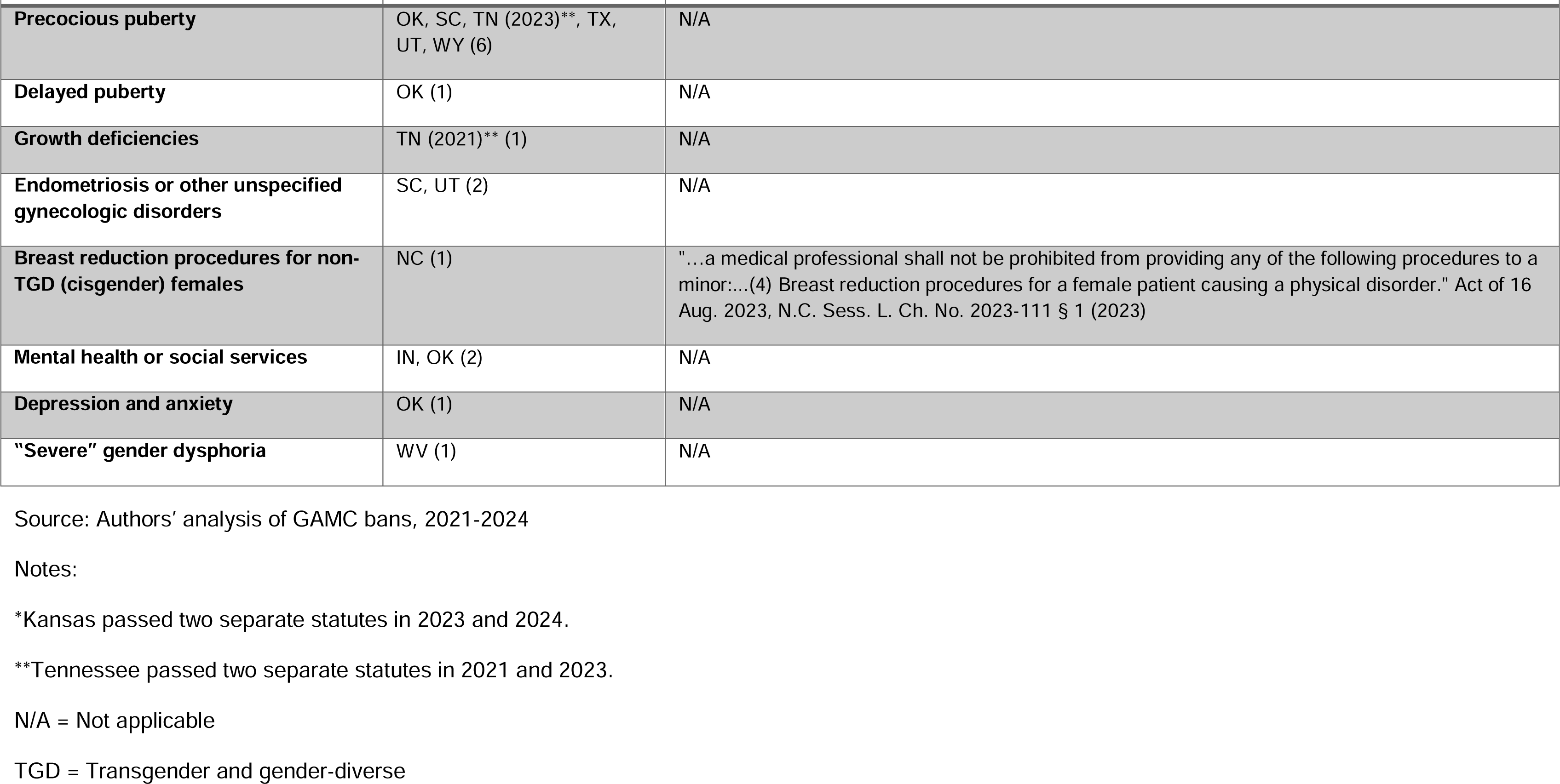
Exceptions in U.S. laws restricting medical care for transgender minors, 2021-2024 (N=30).

| Exception | States with Exception (n) | Example Statutory Language |
| --- | --- | --- |
| **Intersex/variations in sex characteristics** | AL, AZ, AR, FL, GA, ID, IN, IA, KS (2023 & 2024)*, KY, LA, MS, MO, MT, NE, NH, NC, ND, OH, OK, SC, SD, TN (2023)**, TX, UT, WV, WI, WY (29) | "[Restrictions on medical care]...[do] not apply to a procedure undertaken to treat a minor born with a medically verifiable disorder of sex development, including either of the following:<br>(1) An individual born with external biological sex characteristics that are irresolvably ambiguous, including an individual born with 46 XX chromosomes with virilization, 46 XY chromosomes with undervirilization, or having both ovarian and testicular tissue.<br>(2) An individual whom a physician has otherwise diagnosed with a disorder of sexual development, in which the physician has determined through genetic or biochemical testing that a person does not have normal sex chromosome structure, sex hormone production, or sex steroid hormone action for a male or female." (Alabama Vulnerable Child Compassion and Protection Act, Ala. Act No. 2022-289 § 4(b), 2022) |
| **Interventions to revise or address a complication of gender-affirming medical care** | AZ, AR, FL, ID, IN, IA, KS (2024), KY, LA, MS, MO, MT, NE, NC, OH, OK, SC, SD, WV, WI (20) | "[Restrictions on medical care] [do] not prohibit a physician or other practitioner from providing any of the following to a minor:...(3) The treatment of any infection, injury, disease, or disorder that has been caused by or exacerbated by the performance of gender transition procedures." (Act of 5 Apr. 2023, Ind. Pub. L. No. 10, 2023) |
| **Unspecified physical disorders causing imminent danger of death or major bodily impairment** | AZ, AR, FL, ID, IN, IA, LA, MO, NE, NH, NC, OK, SC, TN (2023)**, WV, WI (16) | "The provisions of [restricted medical care] shall not apply to the following:...(4) Any procedure undertaken because the individual suffers from a physical disorder, physical injury, or physical illness that would, as certified by a physician, place the person in imminent danger of death or impairment of major bodily function unless surgery is performed." (Missouri Save Adolescents from Experimentation (SAFE) Act, Mo. S. Sub. No. 2 § A, 2023) |
| **Medically necessary reasons other than gender dysphoria or incongruence** | GA, ID, NC, SC, TN (2021 & 2023)**, UT (7) | "Any surgery, including those listed in [the section on restricted medical care], which a treating physician certifies is medically necessary to treat a physiological condition." (Act of 16 Aug. 2023, N.C. Sess. L. Ch. No. 2023-111 § 1, 2023) |
| **Penile circumcision** | AL, MS, NH, ND (4) | N/A |
| **Removal of a cancerous or diseased body part** | NH, UT (2) | N/A |
| **Prostate cancer** | SC (1) | N/A |
| **Breast cancer** | SC (1) | N/A |
| **Sex-hormone stimulated cancer** | UT (1) | N/A |
| **Partial androgen insensitivity syndrome** | GA (1) | N/A |
| **Precocious puberty** | OK, SC, TN (2023)**, TX, UT, WY (6) | N/A |
| **Delayed puberty** | OK (1) | N/A |
| **Growth deficiencies** | TN (2021)** (1) | N/A |
| **Endometriosis or other unspecified gynecologic disorders** | SC, UT (2) | N/A |
| **Breast reduction procedures for non-TGD (cisgender) females** | NC (1) | "...a medical professional shall not be prohibited from providing any of the following procedures to a minor:...(4) Breast reduction procedures for a female patient causing a physical disorder." Act of 16 Aug. 2023, N.C. Sess. L. Ch. No. 2023-111 § 1 (2023) |
| **Mental health or social services** | IN, OK (2) | N/A |
| **Depression and anxiety** | OK (1) | N/A |
| **“Severe” gender dysphoria** | WV (1) | N/A |

Source: Authors' analysis of GAMC bans, 2021-2024
Notes:
\*Kansas passed two separate statutes in 2023 and 2024.
\*\*Tennessee passed two separate statutes in 2021 and 2023.
N/A = Not applicable
TGD = Transgender and gender-diverse

### Appendix Box 1. Detailed methodology.

We searched for statutes across four full legislative sessions (January 1, 2021, to December 31, 2024) that restrict healthcare services for TGD individuals in all 50 U.S. states, the District of Columbia, U.S. territories, and tribal and federal jurisdictions. Using publicly available legislative trackers,(30-33) one statute of interest was reviewed for each legislative session (AR in 2021, AL in 2022, GA in 2023, OH in 2024) to identify key terms. Lexis Nexis was then queried for all jurisdictions of interest (excepting American Samoa, for which data were not available) with bill text of enrolled and enacted statutes containing the terms: “gender transition,” “gender dysphoria,” “gender reassignment,” “sex reassignment,” “gender-related condition,” “nonambiguous genitalia,” or “irresolvably ambiguous.” Identified statutes meeting our inclusion criteria (Appendix Exhibit A2), including those that were vetoed or under injunction, were cross-validated with the jurisdiction’s government website. They were then assessed through sequential, independent review by two authors (SAM, HCW). For each statute, the authors identified descriptions of sex, gender, and GAMC; restricted healthcare services; stated purpose(s) for which services are restricted; exceptions to restrictions; and penalties for statutory violations. All quoted text denotes statutory language.

**Appendix Figure 1.**
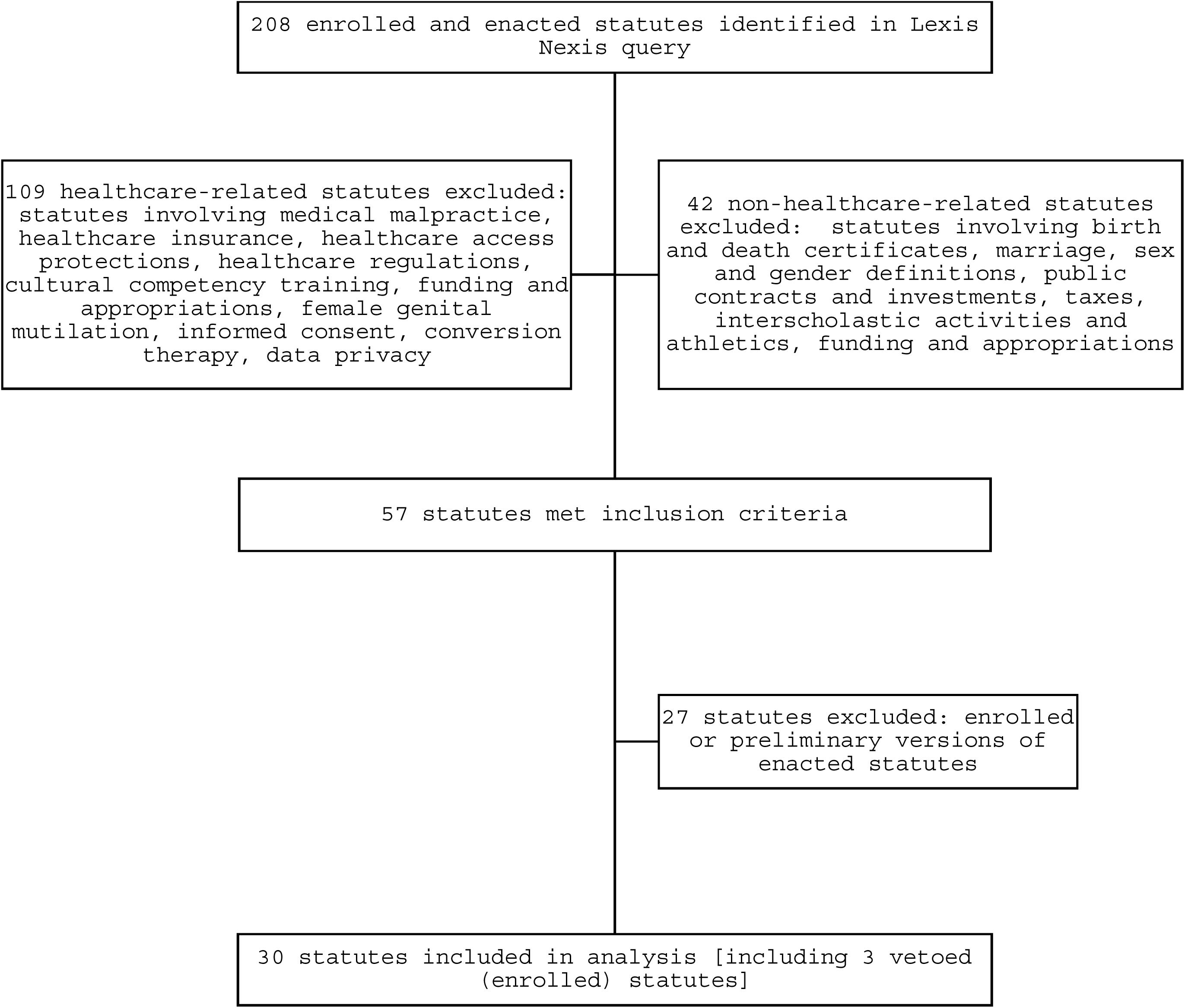
Flow diagram for identification of statutes, inclusion and exclusion criteria.

**Appendix Table 4.**
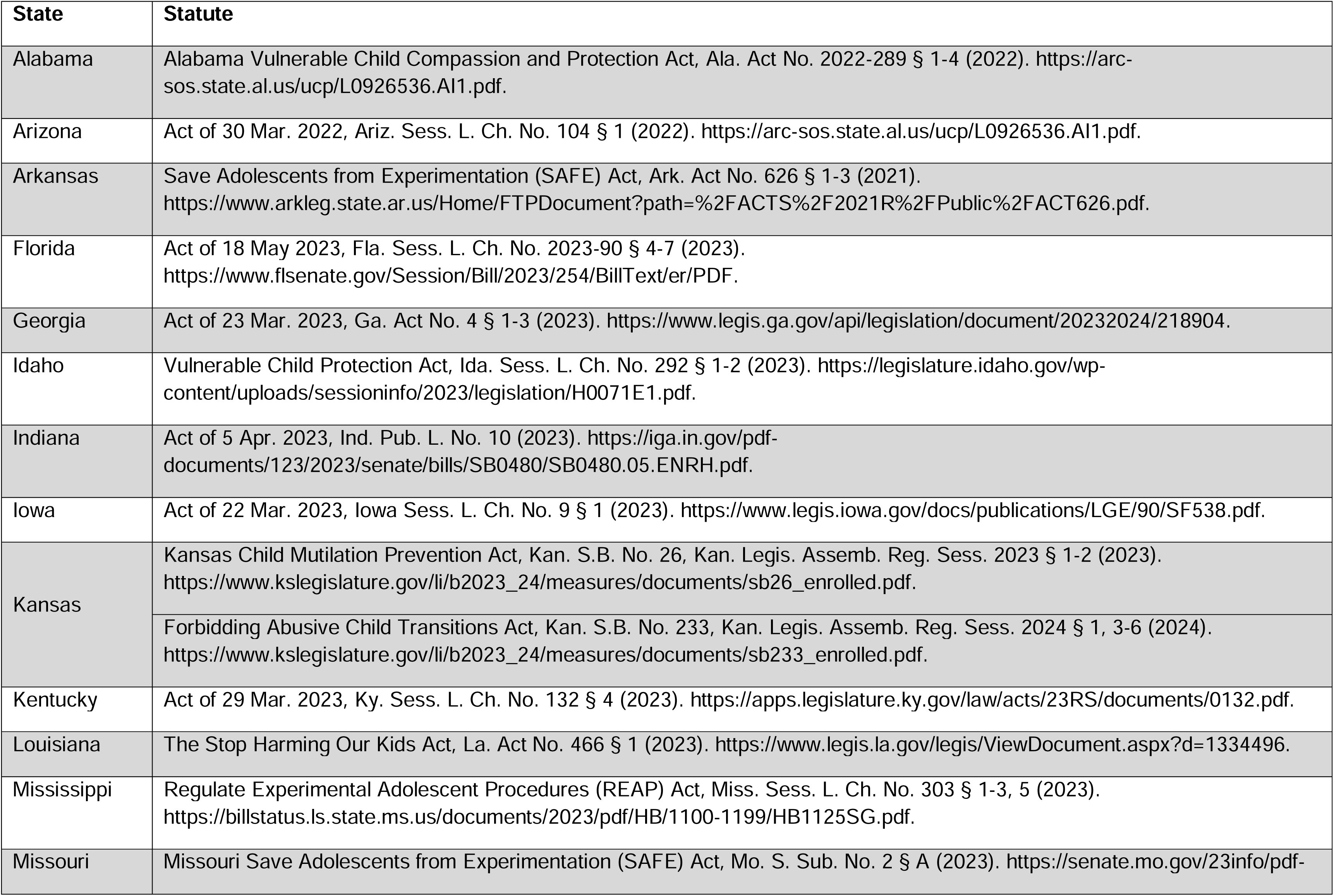
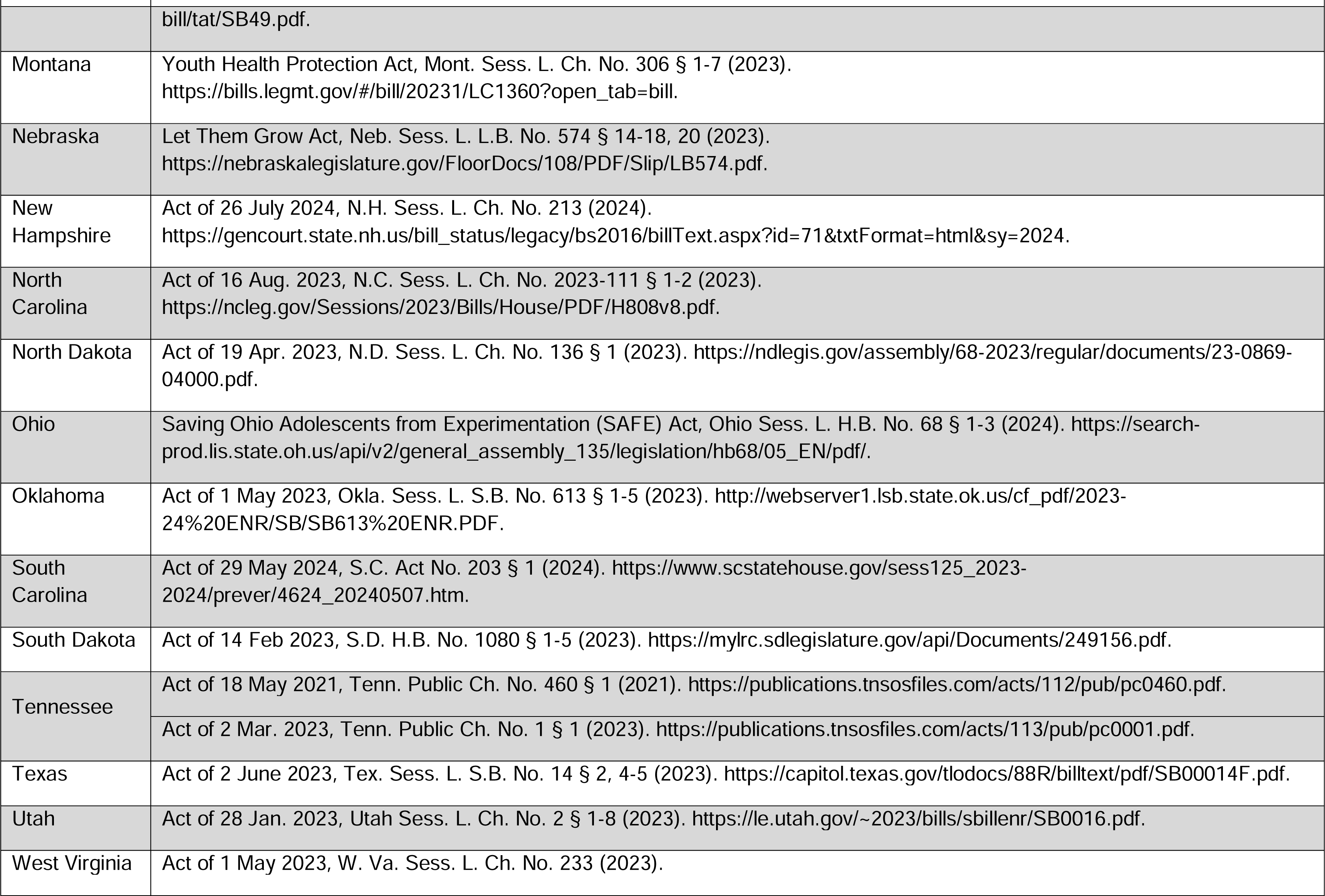
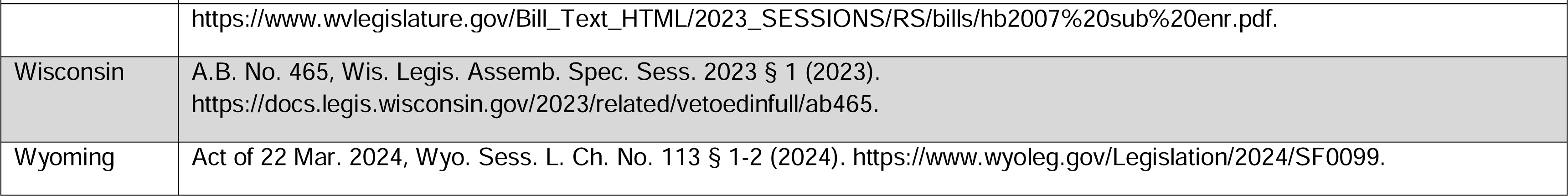
U.S. laws restricting medical care for transgender minors, 2021-2024 (N=30).

## ENDNOTES/REFERENCES

1. Committee on Measuring Sex, Gender Identity, and Sexual Orientation, Committee on National Statistics, Division of Behavioral and Social Sciences and Education, National Academies of Sciences, Engineering, and Medicine. Measuring Sex, Gender Identity, and Sexual Orientation [Internet]. Bates N, Chin M, Becker T, editors. Washington, D.C.: National Academies Press; 2022 [cited 2024 Nov 12]. Available from: https://www.nap.edu/catalog/26424

2. Beischel WJ, Schudson ZC, Hoskin RA, Van Anders SM. The gender/sex 3×3: Measuring and categorizing gender/sex beyond binaries. Psychology of Sexual Orientation and Gender Diversity. 2023 Sep;10(3):355–72.

3. Van Anders SM, Schudson ZC, Abed EC, Beischel WJ, Dibble ER, Gunther OD, et al. Biological Sex, Gender, and Public Policy. Policy Insights from the Behavioral and Brain Sciences. 2017 Oct;4(2):194–201.

4. Richardson SS. Sex Contextualism. Philosophy, Theory, and Practice in Biology [Internet]. 2022 Jan 25 [cited 2024 Dec 8];14(0). Available from: https://journals.publishing.umich.edu/ptpbio/article/id/2096/

5. Krieger N. Genders, sexes, and health: what are the connections—and why does it matter? International Journal of Epidemiology. 2003 Aug;32(4):652–7.

6. Carpenter M. Intersex human rights, sexual orientation, gender identity, sex characteristics and the Yogyakarta Principles plus 10. Culture, Health & Sexuality. 2021 Apr 1;23(4):516–32.

7. Human Rights Violations Against Intersex People: A Background Note. 2019. Office of the High Commissioner for Human Rights. Accessed on 11 Feb 2025. https://www.ohchr.org/sites/default/files/Documents/Issues/Discrimination/LGBT/BackgroundNoteHumanRightsViolationsagainstIntersexPeople.pdf.

8. Blackless M, Charuvastra A, Derryck A, Fausto-Sterling A, Lauzanne K, Lee E. How sexually dimorphic are we? Review and synthesis. Am J Hum Biol. 2000 Mar;12(2):151–66.

9. Abualsaud D, Hashem M, AlHashem A, Alkuraya FS. Survey of disorders of sex development in a large cohort of patients with diverse Mendelian phenotypes. American J of Med Genetics Pt A. 2021 Sep;185(9):2789–800.

10. Hull CL, Fausto-Sterling A. Letter to the Editor. American J Hum Biol. 2003 Jan;15(1):112–6.

11. Sax L. How common is lntersex? A response to Anne Fausto-Sterling. The Journal of Sex Research. 2002 Aug 1;39(3):174–8.

12. Délot EC, Vilain E. Towards improved genetic diagnosis of human differences of sex development. Nat Rev Genet. 2021 Sep;22(9):588–602.

13. Muschialli L, Allen CL, Boy-Mena E, Malik A, Pallitto C, Nihlén Å, et al. Perspectives on conducting “sex-normalising” intersex surgeries conducted in infancy: A systematic review. Siddiq Shekhani S, editor. PLOS Glob Public Health. 2024 Aug 28;4(8):e0003568.

14. Dreger A. Intersex in the age of ethics. Hagerstown, MD: University Publishing Group; 1999.

15. Eder S. How the clinic made gender: the medical history of a transformative idea. Chicago: The University of Chicago Press; 2022. 334 p.

16. Liao LM. Variations in sex development: medicine, culture and psychological practice. Cambridge, United Kingdom; New York, NY: Cambridge University Press; 2023. 1 p.

17. Liao LM, Baratz A. Medicalization of intersex and resistance: a commentary on Conway. Int J Impot Res. 2023 Feb;35(1):51–5.

18. For example, salt-wasting congenital adrenal hyperplasia causes mineralocorticoid deficiency (a physical health risk requiring urgent medical intervention) and may be associated with benign or low-risk VSCs (anatomical characteristics that do not necessitate urgent or otherwise time-sensitive “appearance”-normalizing intervention).

19. The Brussels Collaboration on Bodily Integrity. Genital Modifications in Prepubescent Minors: When May Clinicians Ethically Proceed? The American Journal of Bioethics. 2024 Jul 17;1–50.

20. The Brussels Collaboration on Bodily Integrity. Medically Unnecessary Genital Cutting and the Rights of the Child: Moving Toward Consensus. The American Journal of Bioethics. 2019 Oct 3;19(10):17–28.

21. Human Rights Watch, editor. “I want to be like nature made me” - medically unnecessary surgeries on intersex children in the US. New York, NY: Human Rights Watch; 2017. 179 p.

22. Gender identity may also be understood as an individual’s deeply ingrained sense of being a girl or woman, boy or man, or potentially another gender.

23. Carpenter M, Dalke KB, Earp BD. Endosex. J Med Ethics. 2023 Mar;49(3):225–6.

24. Herman J, Flores A, O’Neill K. How Many Adults and Youth Identify as Transgender in the United States? [Internet]. UCLA School of Law, Williams Institute; 2022. Available from: https://williamsinstitute.law.ucla.edu/wp-content/uploads/Trans-Pop-Update-Jun-2022.pdf

25. Jones, J.M. LGBTQ+ Identification in U.S. Rises to 9.3%. Gallup. 20 Feb 2025. Accessed on 24 Mar 2025. https://news.gallup.com/poll/656708/lgbtq-identification-rises.aspx.

26. For example, surgical interventions that modify chest size or contour are sought to address felt discrepancies between physical embodiment and gender identity among both TGD and non-TGD individuals. This includes mastectomy for transgender men and post-mastectomy reconstructive surgery for non-TGD women.

27. Schall TE, Moses JD. Gender-Affirming Care for Cisgender People. Hastings Center Report. 2023 May;53(3):15–24.

28. Grimstad F, Kremens J, Boskey ER, Wenger H. How Should Clinicians Navigate Decision Making About Genital Reconstructive Surgeries Among Intersex and Transgender Populations? AMA Journal of Ethics. 2023 Jun 1;25(6):E437–445.

29. Rosenthal, G. Samantha. Gender-affirming care has a long history in the US - and not just for transgender people. The Conversation. 27 Mar 2023. Accessed on 17 Dec 2024. https://theconversation.com/gender-affirming-care-has-a-long-history-in-the-us-and-not-just-for-transgender-people-201752.

30. Dawson L, Kates J. Policy Tracker: Youth Access to Gender Affirming Care and State Policy Restrictions. KFF. 19 Mar 2025. Accessed on 24 Mar 2025. https://www.kff.org/other/dashboard/gender-affirming-care-policy-tracker/?utm_campaign=KFF-Global-Health-Policy.

31. Mapping Attacks on LGBTQ Rights in U.S. State Legislatures in 2023. ACLU. 2023. Accessed on 17 Dec 2024. https://www.aclu.org/legislative-attacks-on-lgbtq-rights-2023. ACLU;

32. Mapping Attacks on LGBTQ Rights in U.S. State Legislatures in 2024. ACLU. 2024. Accessed on 17 Dec 2024. https://www.aclu.org/legislative-attacks-on-lgbtq-rights-2024.

33. Equality Maps: Bans on Best Practice Medical Care for Transgender Youth. Movement Advancement Project. Accessed on 24 Mar 2025. https://www.lgbtmap.org/equality-maps/healthcare_youth_medical_care_bans.

34. Lau H, Fedders B. Scrutinizing Transgender Healthcare Bans Through Intersex Exceptions [Internet]. 2024 [cited 2024 Nov 12]. Available from: https://www.ssrn.com/abstract=4935674

35. Mapping the Intersex Exceptions: Anti-Trans Legislation Across the United States Permits Rights Violations Against Intersex Children. Human Rights Watch. 2022. Accessed on 17 Dec 2024. https://www.hrw.org/feature/2022/10/26/mapping-the-intersex-exceptions?gad_source=1&gclid=Cj0KCQiAuou6BhDhARIsAIfgrn5u52MVsF4fALatBpOn0hFIKx82NMGnsl6yuumVSDtzaK5XioggBkEaAqiMEALw_wcB.

36. See Appendix.

37. A GAMC ban was passed in Kansas in 2023 and 2024 (both vetoed) and Tennessee in 2021 and 2023.

38. In five states (AR, KY, LA, NC, OH), a GAMC ban was enacted after a veto override.

39. Act of 18 May 2021, Tenn. Public Ch. No. 460 § 1 (2021). https://publications.tnsosfiles.com/acts/112/pub/pc0460.pdf.

40. Act of 2 Mar. 2023, Tenn. Public Ch. No. 1 § 1 (2023). https://publications.tnsosfiles.com/acts/113/pub/pc0001.pdf.

41. Waldeck S. Using Male Circumcision to Understand Social Norms as Multipliers. University of Cincinnati Law Review. 2007 Oct 28;72.

42. Act of 26 July 2024, N.H. Sess. L. Ch. No. 213 (2024). https://gencourt.state.nh.us/bill_status/legacy/bs2016/billText.aspx?id=71&txtFormat=html&sy=2024.

43. Walden RL, Abdulcadir J, Earp BD. Labiaplasty in Minors: Medicalizing Mutilation? Arch Sex Behav. 2025 Jan;54(1):95–106.

44. Luchristt D, Sheyn D, Bretschneider CE. National Estimates of Labiaplasty Performance in the United States From 2016 to 2019. Obstetrics & Gynecology. 2022 Aug;140(2):271–4.

45. Kalampalikis A, Michala L. Cosmetic labiaplasty on minors: a review of current trends and evidence. Int J Impot Res. 2023 May;35(3):192–5.

46. Dai D, Charlton BM, Boskey ER, Hughes LD, Hughto JMW, Orav EJ, et al. Prevalence of Gender-Affirming Surgical Procedures Among Minors and Adults in the US. JAMA Netw Open. 2024 Jun 27;7(6):e2418814.

47. 2023 ASPS Procedural Statistics Release. Plastic & Reconstructive Surgery. 2024 Sep;154(3S):1–41.

48. American Psychiatric Association, editor. Diagnostic and statistical manual of mental disorders: DSM-5-TR^TM^. Fifth edition, text revision. Washington, DC: American Psychiatric Association Publishing; 2022. 1050 p.

49. Although it might be conceded that most intersex “normalization” procedures, like most circumcisions, are not strictly necessary on grounds of physical health, it could still be argued that these procedures are intended to promote the child’s psychological well-being, due to an expectation the child will feel happier in a body that conforms to what is typical for someone of their anticipated gender identity (where this, in turn, is assumed to correspond to their sex as recorded at birth). However, such a justification would fail to explain, within state-level bans on GAMC, the simultaneous exception for circumcision and restriction on voluntary GAMC for TGD individuals, who, in the latter case, similarly argue that such interventions will make them feel happier or more “at home” in their bodies.

50. Reis E. Intersex Surgeries, Circumcision, and the Making of “Normal.” In: Denniston GC, Hodges FM, Milos MF, editors. Genital Cutting: Protecting Children from Medical, Cultural, and Religious Infringements [Internet]. Dordrecht: Springer Netherlands; 2013 [cited 2024 Jun 13]. p. 137–47. Available from: https://link.springer.com/10.1007/978-94-007-6407-1_10

51. Cass, H. 2024. Independent review of gender identity services for children and young people: Final report. NHS England. Accessed on 11 Feb 2025. https://cass.independent-review.uk/home/publications/final-report/.

52. Grijseels DM. Biological and psychosocial evidence in the Cass Review: a critical commentary. International Journal of Transgender Health. 2024 Jun 8;1–11.

53. Horton C. The Cass Review: Cis-supremacy in the UK’s approach to healthcare for trans children. International Journal of Transgender Health. 2024 Mar 14;1–25.

54. Noone C, Southgate A, Ashman A, Quinn É, Comer D, Shrewsbury D, et al. Critically Appraising the Cass Report: Methodological Flaws and Unsupported Claims [Internet]. 2024 [cited 2025 Feb 11]. Available from: https://osf.io/uhndk

55. Supporters of the bans might respond that, even if “meaningful participation” is possible, TGD minors are too young to understand the implications of GAMC. In the first place, such an argument would be inconsistent with standard approaches to pediatric ethics, whereby parents and healthcare professionals, not legislators, are expected to evaluate a minor’s understanding of a proposed intervention, and parental “proxy” permission, along with the assent of the minor, are usually considered sufficient for authorization. But even if such an argument were granted, it would fail to explain the exceptions for anatomically similar “normalizing” procedures on intersex infants and children, given that these are typically carried out when the child has considerably less, or no, understanding of what is at stake.

56. Giordano S, Garland F, Holm S. Gender dysphoria in adolescents: can adolescents or parents give valid consent to puberty blockers? J Med Ethics. 2021 Mar 10;medethics-2020-106999.

57. Svoboda JS. Promoting genital autonomy by exploring commonalities between male, female, intersex, and cosmetic female genital cutting. Global Discourse. 2013 Jun;3(2):237–55.

58. DeLaet DL. Genital Autonomy, Children’s Rights, and Competing Rights Claims in International Human Rights Law. Int J Child Rights. 2012;20(4):554–83.

59. Exceptional circumstances include ones in which a physical health emergency must be addressed while the individual lacks capacity.

60. Kraus C, Phan-Hug F, Ansermet F, Meyrat BJ. Améliorer les pratiques de soins pour les personnes présentant une variation du développement du sexe en Suisse. L’École de Lausanne (depuis 2005). droitcultures [Internet]. 2021 Jan 26 [cited 2025 Feb 4];(80). Available from: http://journals.openedition.org/droitcultures/6610

61. Phan-Hug F, Kraus C, Paoloni-Giacobino A, Fellmann F, Typaldou SA, Ansermet F, et al. [Patients with variations of sex development : an example of interdisciplinary care]. Rev Med Suisse. 2016 Nov 9;12(538):1923–9.

62. Moran ME, Karkazis K. Developing a Multidisciplinary Team for Disorders of Sex Development: Planning, Implementation, and Operation Tools for Care Providers. Advances in Urology. 2012;2012:1–12.

63. Karkazis K, Tamar-Mattis A, Kon AA. Genital Surgery for Disorders of Sex Development: Implementing a Shared Decision-Making Approach. Journal of Pediatric Endocrinology and Metabolism [Internet]. 2010 Jan [cited 2025 Feb 4];23(8). Available from: https://www.degruyter.com/document/doi/10.1515/jpem.2010.129/html

64. Model Legislation: The JUST FACTs Act (The JUSTice For Adolescent and Child Transitioners Act). Do No Harm. 2023. Accessed on 17 Dec 2024. https://donoharmmedicine.org/wp-content/uploads/2023/01/Do-No-Harm-The-JUST-FACTs-Act-Model-Legislation.pdf.

65. Assemb. Bill No. 977. Wis. Legis. Assemb. LRB-0396 (2021). https://docs.legis.wisconsin.gov/2021/related/drafting_files/assembly_intro_legislation/assembly_bills_not_enacted/2021_ab_0977/01_ab_977/21_0396df.pdf. Cf. Help Not Harm - Model Bill Language, Version 2.0, Family Policy Alliance.

66. A.B. No. 465, Wis. Legis. Assemb. Spec. Sess. 2023 § 1 (2023). https://docs.legis.wisconsin.gov/2023/related/vetoedinfull/ab465.

67. Beshear, A. Veto Message, Ky. S.B. No. 150 (2023). https://apps.legislature.ky.gov/record/23rs/sb150/veto.pdf.

68. Cooper, R. Veto Message, N.C. H.B. No. 808 (2023). https://webservices.ncleg.gov/ViewBillDocument/2023/6811/0/H808-BD-NBC-11125.

69. DeWine, M. Veto Message, Ohio. H.B. No. 68 (2023). https://www.legislature.ohio.gov/assets/legislation/legislation-documents/135/VetoMessageSUBHB68.pdf.

70. Edwards, JB. Veto Message, La. H.B. No. 648 (2023). https://gov.louisiana.gov/assets/2023Vetoes/HB648.pdf.

71. Kelly, L. Veto Message, Kan. S.B. No. 26 (2023). https://www.sos.ks.gov/publications/sessionlaws/2023/Message-07-SB-26.html.

72. Kelly, L. Veto Message, Kan. S.B. No. 233 (2024). https://www.sos.ks.gov/publications/sessionlaws/2024/Message-02-SB-233.html.

73. L. W., By and Through Her Parents and Next Friends, Samantha Williams and Brian Williams, et al., Petitioners v. Jonathan Skrmetti, Attorney General and Reporter for Tennessee, et al. Supreme Court of the United States. Accessed on 17 Dec 2024. https://www.supremecourt.gov/Search.aspx?FileName=/docket/docketfiles/html/public/23-466.html.

74. Legal Frameworks: Restrictions on Interventions on Intersex Minors. ILGA World Database. Accessed on 17 Dec 2024. https://database.ilga.org/interventions-intersex-minors.

75. OHCHR Technical Note on the Human Rights of Intersex People: Human Rights Standards and Good Practices. Office of the High Commissioner for Human Rights, 3 Nov 2023. Accessed on 17 Dec 2024. https://www.ohchr.org/sites/default/files/2023-11/ohchr-technical-note-rights-intersex-people.pdf.

76. U.S. Department of Health and Human Services, Office of the Assistant Secretary for Health (2025) Advancing Health Equity for Intersex Individuals. Accessed on 2 Feb 2025. https://web.archive.org/web/20250126102630/https://www.hhs.gov/sites/default/files/intersex-health-equity-report.pdf.

77. HHS supports intersex bodily autonomy in first-ever health equity report. interACT. Accessed on 2 Feb 2025. https://interactadvocates.org/hhs-supports-bodily-autonomy/.

78. Exec. Order No. 14168, 90 Fed. Reg. 8615 (January 20, 2025).

79. Carpenter M. Is It Ever OK to Reclassify Someone Out of Their Birth-Observed Sex Without Personal Consent? How Do We Manage Competing Methods of Classifying Sex? The American Journal of Bioethics. 2024 Nov;24(11):18–20.

80. Trump’s Executive Order Ignores Science to Push Discriminatory Agenda. interACT. Accessed on 9 Feb 2025. https://interactadvocates.org/trumps-executive-order-ignores-science-to-push-discriminatory-agenda/.

81. Trump’s New Policy on “Gender Ideology” and “Biologial Truth.” InterAction Australia. Accessed on 9 Feb 2025. https://interaction.org.au/41622/trumps-new-policy-2025-01/.

82. Exec. Order No. 14187, 90 Fed. Reg. 8771 (Januxary 28, 2025).

83. Case: PFLAG, Inc. v. Donald J. Trump. Civil Rights Litigation Clearinghouse. Accessed on 24 Mar 2025. https://clearinghouse.net/case/46028/.

84. Case: State of Washington v. Department of Justice. Civil Rights Litigation Clearinghouse. Accessed on 24 Mar 2025. https://clearinghouse.net/case/46065/.

85. Case: Kingdom v. Trump. Civil Rights Litigation Clearinghouse. Accessed on 24 Mar 2025. https://clearinghouse.net/case/46207/.

86. Case: Doctors for America v. Office of Personnel Management. Civil Rights Litigation Clearinghouse. Accessed on 24 Mar 2025. https://clearinghouse.net/case/46029/.

87. Dawson L. President Trump’s Executive Order on Gender Affirming Care: Responses by Providers, States, and Litigation. KFF. 11 Feb 2025. Accessed on 24 Mar 2025. https://www.kff.org/policy-watch/president-trumps-executive-order-on-gender-affirming-care-responses-by-providers-states-and-litigation/.

88. 18 U.S.C. § 116.

89. 18 U.S.C. § 116b1.

90. Chase, C. 2002."Cultural practice" or “reconstructive surgery”? U.S. genital cutting, the intersex movement and medical double standards. In James, S. M., & Robertson, C. C. (Eds.). (2002). Genital cutting and transnational sisterhood: Disputing US polemics. University of Illinois Press.

91. Ehrenreich, N., and M. Barr. 2005. Intersex surgery, female genital cutting, and the selective condemnation of cultural practices. Harvard Civil Rights. Civil Liberties Law Review 40 (1):71–140. https://pdfs.semanticscholar.org/a986/deba1d02e1035596bfde5befe171eaa95252.pdf.

92. O’Neill S, Bader D, Kraus C, Godin I, Abdulcadir J, Alexander S. Rethinking the Anti-FGM Zero-Tolerance Policy: from Intellectual Concerns to Empirical Challenges. Curr Sex Health Rep. 2020 Dec;12(4):266–75.

93. Carpenter M. Protecting intersex people from harmful practices in medical settings: a new benchmark in the Australian Capital Territory. Australian Journal of Human Rights. 2023 May 4;29(2):409–17.

94. Bader D, Mottier V. Femonationalism and populist politics: The case of the Swiss ban on female genital mutilation. Nations and Nationalism. 2020 Jul;26(3):644–59.

95. Mendez, Bryce HP. (2025, January 10). FY2025 NDAA: TRICARE Coverage of Gender-Affirming Care. (CRS Report No. IN12401).

96. H.R. 5009 - 118th Congress (2023-2024): The Servicemember Quality of Life Improvement and National Defense Authorization Act for Fiscal Year 2025 § 708. (2024, December 7). https://docs.house.gov/billsthisweek/20241209/RCP_HR5009_xml%5b89%5d.pdf.

97. Kaiser J, Wadman M. Health agencies purge Trump-targeted programs and websites [Internet]. 2025 [cited 2025 Feb 9]. Available from: https://www.science.org/content/article/health-agencies-purge-trump-targeted-programs-and-websites

98. The Help Not Harm Act, Kan. S.B. No. 63 § 1, 3-5 (2025). https://sos.ks.gov/publications/register/Volume-44/Issues/Issue-08/02-20-25-52891.html.

99. Kelly, L. Veto Message, Kan. S.B. No. 63 (2025). https://www.kssos.org/publications/sessionlaws/2025/Message-01-SB-63.html.

100. Walker, J. Legislators pass bill to fully ban gender-affirming care, scrapping exemption for suicidal teens. WV Public Broadcasting. Available from https://wvpublic.org/legislators-pass-bill-to-fully-ban-gender-affirming-care-scrapping-exemption-for-suicidal-teens/

